# The impact of side effect framing on COVID-19 booster vaccine intentions in an Australian sample

**DOI:** 10.1101/2022.05.09.22274840

**Authors:** K. Barnes, K. Faasse, B. Colagiuri

## Abstract

**Objective:** To evaluate the effect of presenting positively attribute-framed side effect information on COVID-19 booster vaccine intention relative to standard negatively-framed wording and a no-intervention control.

**Design setting and participants:** A representative sample of Australian adults (*N*=1,204) were randomised to one of six conditions within a factorial design: Framing (Positive; Negative; Control) * Vaccine (Familiar (Pfizer); Unfamiliar (Moderna)).

**Intervention:** Negative Framing involved presenting the likelihood of experiencing side effects (e.g., heart inflammation is very rare, 1 in every 80,000 will be affected), whereas Positive Framing involved presenting the same information but as the likelihood of *not* experiencing side effects (e.g., 79,999 in every 80,000 will not be affected).

**Primary Outcome:** Booster vaccine intention measured pre- and post-intervention.

**Results:** Positive Framing (*M*=75.7, *SE*=0.9, 95% CI[73.9, 77.4]) increased vaccine intention relative to Negative Framing (*M*=70.7, *SE*=0.9, 95% CI[68.9, 72.4]) overall (*F*(1, 1192)=4.68, *p*=.031, *η*_*p*_^*2*^=.004). Framing interacted with Vaccine and Baseline Intention (*F*(2, 1192)=6.18, *p*=.002, *η*_*p*_^*2*^=.01). Positive Framing was superior, or at least equal, to Negative Framing and Control at increasing Booster Intention, irrespective of the participants pre-intervention level of intent. Side effect worry and perceived severity mediated the effect of Positive vs. Negative Framing across vaccines.

**Conclusion:** Positive framing of side effect information appears superior for increasing vaccine intent relative to the standard negative wording currently used.

**Pre-registration:** See: aspredicted.org/LDX_2ZL

## Introduction

Reasons for vaccine hesitancy are multifaceted^1,2^. However, side effect apprehension is a primary factor^1^, with previous experience of COVID-19 vaccine side effects shown to reduce later booster vaccine intention^3^. To achieve effective societal protection from COVID-19, behavioural intervention is required to reduce apprehension and increase vaccine acceptance^4^.

Side effect information framing has been suggested as such an intervention^5^. Standard negative wording is typically used to present side effect information on official sources such as the Australian Government and AusVaxSafety (a national vaccine safety system) websites. For example, “33% of people ***were affected*** by headaches after their second Pfizer dose”. Our research suggests negative wording of this type may increase hesitancy relative to positive wording (i.e., presenting the number ***not affected***) ^6^. Further, consistent with prior research demonstrating that vaccine relevance or familiarity moderates the effect of framing^7,8^, this effect was especially pronounced when associated with an unfamiliar vaccine^6^.

Updating currently presented side effect information with a positively framed counterpart is both easy to implement and does not violate patient informed consent (given that statistical information is equivalent to the standard negative form) ^9^. As such, framing may be particularly well suited to increasing vaccine acceptance. However, in our previous research conducted in a UK sample, framing was applied to side effect frequency bands presented in manufacturer Patient Information Leaflets (PILs). This statistical information is commonly presented in the EU, but not in Australia.

In the present study, we therefore tested an intervention more similar to current Australian sources of side effect information (such as AusVaxSafety). Information detailing severe side effects (heart inflammation and anaphylaxis^10,11^) and side effect-induced daily disruptions (data from AusVaxSafety) were presented to participants who had received two doses of a COVID-19 vaccine but no booster. This information was presented using the standard negative wording (Negative Framing group), positive wording (Positive Framing group), or not at all (No-Intervention Control group).

Pre-registered hypotheses were as follows: 1) being presented with objective side effect information, irrespective of type of framing, would increase Booster Intention relative to Control; 2) Positive Framing would increase Booster Intention relative to Negative Framing; 3) Framing (Positive or Negative) would interact with Vaccine Familiarity, with the effect of Positive Framing on Booster Intention being stronger for a less familiar vaccine (i.e., Moderna relative to Pfizer: at the time of data collection ∼10,000,000 doses of Moderna had been administered compared to 38,454,860 doses of Pfizer). Secondary-outcomes were explored as potential mediators of the framing effect^9,12,13^ (see Supplementary Materials: S1.1).

## Methods

### Participants

Participants (*N*=1,204) were recruited via Pureprofile, an ISO-certified panel provider. Inclusion criteria were: 1) 18+ years of age; 2) residing in Australia; 3) self-reported English fluency; 4) two doses of a COVID-19 vaccine; 5) no booster vaccine. Data were collected between 3^rd^–13^th^ December 2021, just prior to the Omicron outbreak in Australia. Participants received $2 for a 10-minute survey. All procedures in this pre-registered study (aspredicted.org/LDX_2ZL) were approved by the University of Sydney Human Research Ethics Committee (reference, 2021/871), and all participants provided informed consent. Reporting is consistent with STROBE guidelines (Supporting Information: S1.2).

### Data Collection

Data were collected on Pureprofile’s inhouse platform. Stratification and randomisation (via random number generation) occurred via inbuilt code. Quotas were set for a minimum *N*=200 in each experimental condition (details below). All items required a response before advancement. Several additional items concerning COVID-19 were collected prior to randomisation for a separate pre-registered study (aspredicted.org/XSS_ZD1).

### Design

Participants were stratified by their previous two COVID-19 vaccine doses (2xPfizer/2xAstraZeneca/Combination) and randomised to one of six experimental conditions in a 3 (Framing: Positive, Negative, Control) by 2 (Vaccine: Familiar(Pfizer) vs. Unfamiliar(Moderna)) factorial design.

The primary outcome was COVID-19 Booster Intention post-intervention for the assigned vaccine (either Pfizer or Moderna). The following secondary outcomes were measured to test for mediation of the framing effect: 1) Side Effect Worry; 2) Side Effect Severity; 3) Booster Protection. Two additional outcomes were measured. The first was Booster Intention for the “unassigned” vaccine (for the Framing groups, this was the vaccine for which they received no side-effect information; for the Control group, this was simply the other vaccine). This was used to explore possible generalisation of the framing effect to the other vaccine. The second was familiarity with the side effects of the Pfizer and Moderna vaccines measured pre-intervention. This was used to confirm that participants’ responses reflected the assumed Familiarity with each vaccine (i.e., Familiar/Pfizer>Unfamiliar/Moderna). All measures were rated on 100-point Visual Analogue Scale (VAS).

### Framing Intervention

The intervention occurred in five stages. In stage 1, participants selected whether ‘daily disruptions’ or ‘serious side effects’ were their primary vaccination concern. In stage 2, they estimated the percentage of the population they believed would experience six side effects common to both the Pfizer and Moderna vaccine (i.e., local reaction, fatigue, headache, muscle or joint pain, gastrointestinal symptoms, fever) after receiving the framed vaccine. In stage 3, they were shown their responses from stage 2 against veridical percentages from Australian population data (derived from AusVaxSafety; 8^th^ November 2021) for 1 minute. Stages 2 and 3 were performed in order to encourage participants to engage with and process the presented side effect information. The primary components of the intervention occurred in stages 4 and 5. In stage 4, participants viewed framed information (either positive or negative depending on group assignment) in the form of infographics regarding ‘daily disruptions’ from side effects. In stage 5 this was also done regarding ‘serious side effects’ (estimates from^10,11^). Stages 4 and 5 were displayed for 30 seconds each. The exact order of presentation for stages 4 and 5 depended on the participant’s primary concern from stage 1. For example, if they indicated that they were more concerned with ‘daily disruptions’ then that information was first presented at stage 4.

The information presented to the Positive and Negative framing groups differed only in terms of whether this information stated the likelihood of experiencing or *not* experiencing the described side effect. Those assigned to the Negative Framing group saw ‘standard’ wording regarding side effects (e.g., Heart Inflammation: occurrence is very rare, 1 in every 80,000 will be affected), while those in the Positive Framing group saw the logical inverse (e.g., occurrence is very rare, 79,999 in every 80,000 *will* ***not be affected***). Example infographics are displayed in Figure 1 (all infographics presented in S1.4). Framed information was not presented to the Control group. Instead, participants undertook an activity of their choosing for an equivalent intervention duration (2 minutes).

**Figure 1:**
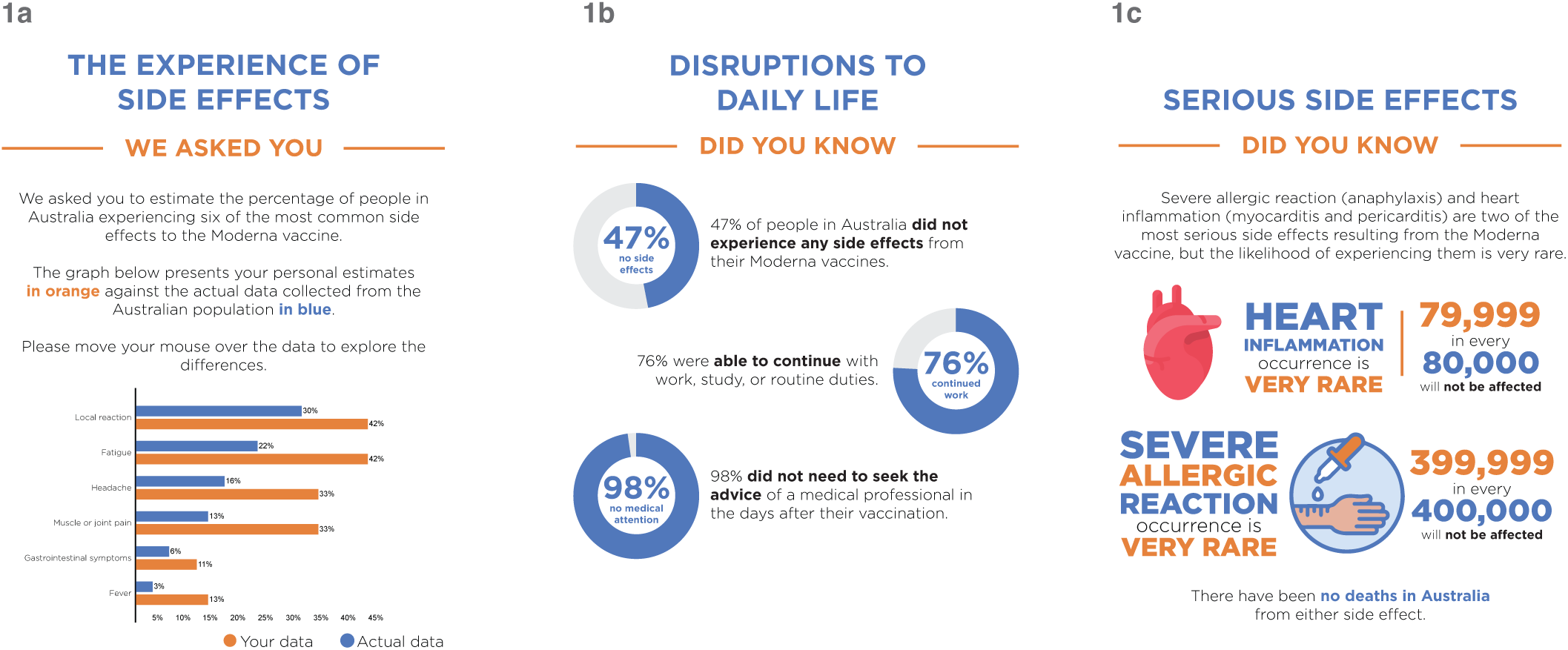
Example materials used to present side effect information to those who received Positive Framing of the Unfamiliar vaccine (i.e., Moderna). 1a demonstrates the wording when presenting estimated and actual prevalence rates for the six common side effects, 1b concerns daily disruptions, and 1c serious adverse events from vaccination.

### Procedure

Prior to randomisation, all participants provided demographic information, as well as vaccine and COVID-19 history (see Supporting Information (S1.3) for survey items). They also completed baseline measures for the primary (Booster Intention) and secondary outcomes, before being assigned to a condition. Please note, Baseline Intention was collected for both vaccines (Pfizer and Moderna). Dependent on randomisation, one of these ratings became the baseline for the framed vaccine, and the other the ‘unassigned’ vaccine (i.e., to explore generalisation).

Following group assignment, framing information (positive or negative) regarding side-effect likelihood for the assigned vaccine was presented to those in the intervention groups. All participants subsequently provided post-manipulation ratings for primary and secondary outcomes. See Figure 2 for a visual representation of the Procedure.

**Figure 2:**
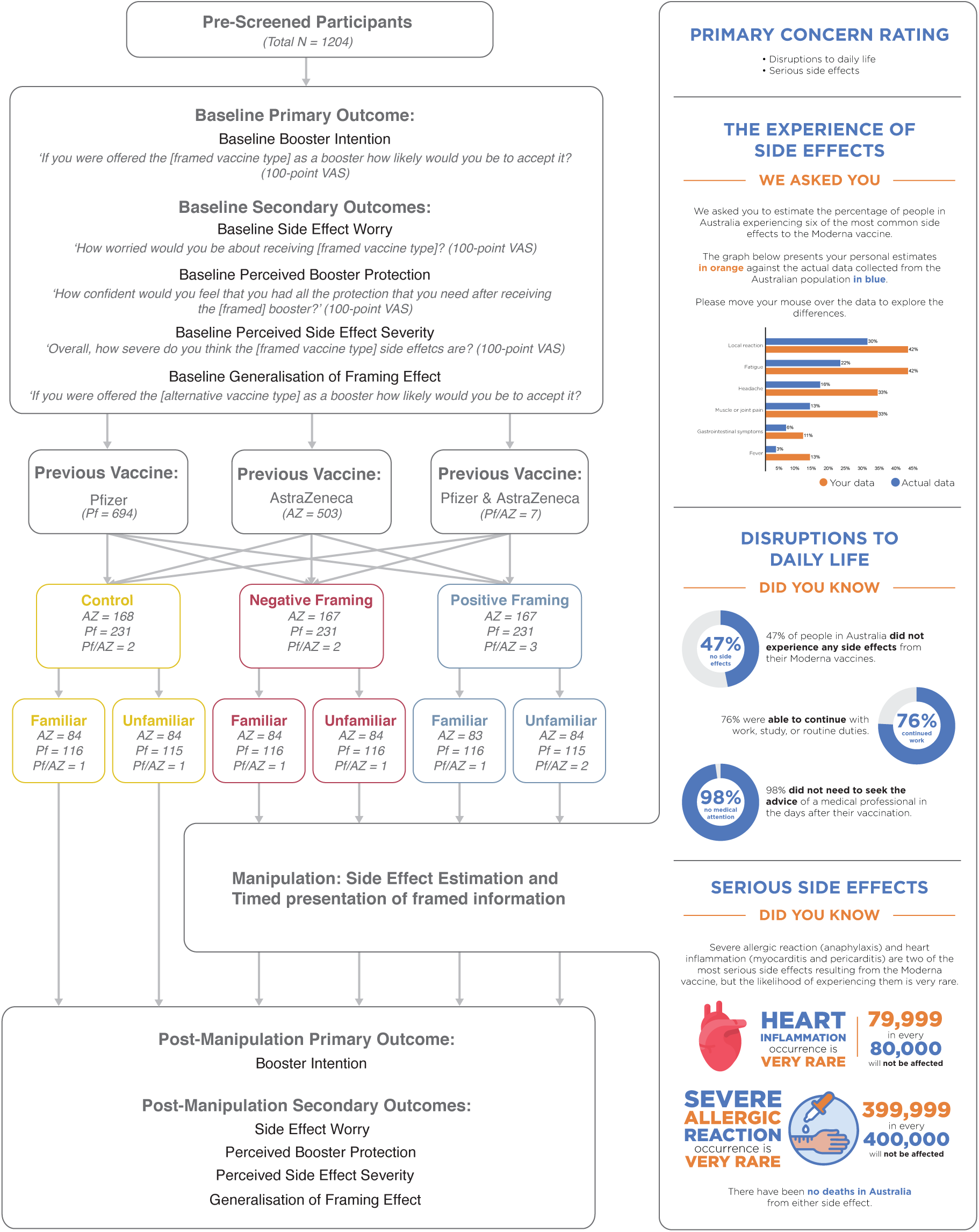
Sample size, item wording, and experimental design (example: Unfamiliar/Positive Frame). Participants provided Baseline Booster Intention ratings for both vaccines (Pfizer and Moderna). Dependent on randomisation, one became the baseline for the framed vaccine and the other to explore generalisation of the framing effect (i.e., to unassigned vaccine). Participants were stratified by previous vaccine and randomised to one of six groups (2*3 factorial design). Those receiving Positive or Negative Framing underwent a manipulation with five stages: rating of primary concerns; active estimation of side effects; presentation of estimated side effects against population data; framed information regarding daily disruptions; framed information regarding serious side effects. Pre-registered analysis of all secondary outcomes are presented as Supporting Information (S1.1).

### Analysis and Sample

Statistical analysis was performed using R (version 4.1.1). Primary analysis was a 3(Framing)*2(Vaccine) ANCOVA on post-intervention Booster Intention. Pre-specified orthogonal contrasts were: Contrast1 (Control vs. Framing [Positive and Negative combined]); Contrast2 (Positive vs. Negative). Differences in baseline Booster Intention (the covariate) were observed with Vaccine Familiarity (*F*(1, 1198)=172.49, *p*<.0001, *η*_*p*_^*2*^=.13). As such, interactions between the covariate and manipulated variables (i.e., Framing and Familiarity) were included in the model, as recommended^14,15^. Mediation using the Lavaan package was performed using the secondary outcomes (Side Effect Worry; Side Effect Severity; Booster Protection).

Sample size was calculated via an *a priori* power analysis (95% power, alpha=.05, effect size *f*^*2*^=0.02) for a separate concurrent study run that contained more predictors (*N*=9), and therefore required more power (see study pre-registration).

## Results

### Sample

A total of 2,639 participants expressed interest. Of these, 998 did not meet inclusion criteria, 3 terminated at consent, 296 withdrew before completion, and 138 were excluded on pre-registered quality-control criteria (see S1.5). The final sample included 1,204 participants.

### Demographics

Demographic information, COVID-19 exposure and vaccination history can be found in Table 1. Participant frequency by SA4 postal region is presented in Figure 3.

**Table 1:** Sample descriptive statistics presented against population data generated from the Australian Bureau of Statistics, Education and Work Dataset 2021. *Nb*: Education sums to 109% due to calculation relying on separate ISCED and ASCED items to calculates estimates for Primary and High School completers. Dashes denote unavailable population data.

|  | Control<br>Familiar<br>*N=201* | Control<br>Unfamiliar<br>*N=200* | Negative<br>Familiar<br>*N=201* | Negative<br>Unfamiliar<br>*N=201* | Positive<br>Familiar<br>*N=200* | Positive<br>Unfamiliar<br>*N=201* | All<br>Respondents<br>(%) | Australian<br>population<br>(%) |
| --- | --- | --- | --- | --- | --- | --- | --- | --- |
| **Demographic Information** |  |  |  |  |  |  |  |  |
| **State/territory** |  |  |  |  |  |  |  |  |
| New South Wales | 68 | 64 | 62 | 67 | 65 | 53 | 31.5 | 31.7 |
| Victoria | 48 | 48 | 58 | 52 | 54 | 57 | 26.3 | 26.3 |
| Queensland | 42 | 42 | 36 | 42 | 37 | 46 | 20.3 | 20.0 |
| Western Australia | 21 | 17 | 16 | 19 | 21 | 27 | 10.0 | 10.4 |
| South Australia | 12 | 23 | 16 | 11 | 18 | 10 | 7.5 | 6.9 |
| Tasmania | 5 | 0 | 8 | 4 | 6 | 3 | 2.2 | 2.1 |
| Australian Capital Territory | 5 | 4 | 4 | 5 | 2 | 2 | 1.8 | 1.7 |
| Northern Territory | 0 | 2 | 1 | 1 | 0 | 0 | 0.3 | 0.8 |
| **Region** |  |  |  |  |  |  |  |  |
| Metro | 153 | 147 | 147 | 147 | 129 | 140 | 71.7 | 67.7 |
| Regional | 48 | 53 | 54 | 54 | 71 | 61 | 28.3 | 32.3 |
| **Employment** |  |  |  |  |  |  |  |  |
| Employed full-time | 85 | 79 | 88 | 88 | 78 | 82 | 41.5 | 49.4 |
| Employed part-time | 40 | 47 | 35 | 36 | 35 | 31 | 18.6 | 21.7 |
| Self employed | 6 | 9 | 5 | 9 | 9 | 8 | 3.8 | - |
| Unemployed (looking) | 9 | 9 | 10 | 13 | 7 | 14 | 5.1 | 1.3 |
| Carer | 12 | 10 | 7 | 12 | 14 | 13 | 5.6 | - |
| Student | 4 | 9 | 9 | 6 | 9 | 6 | 3.6 | - |
| Retired | 44 | 37 | 45 | 37 | 47 | 46 | 21.3 | - |
| Other | 1 | 0 | 2 | 0 | 1 | 1 | 0.4 | - |
| **Education** |  |  |  |  |  |  |  |  |
| Primary school | 1 | 0 | 0 | 1 | 2 | 0 | 0.3 | 4.5 |
| High school | 51 | 58 | 46 | 53 | 57 | 60 | 27.0 | 45.2 |
| Technical certificate | 28 | 30 | 34 | 29 | 29 | 34 | 15.3 | 18.5 |
| Advanced diploma/diploma | 25 | 26 | 41 | 20 | 25 | 23 | 13.3 | 10.1 |
| Bachelor's degree | 59 | 40 | 48 | 59 | 51 | 49 | 25.4 | 20.6 |
| Graduate diploma/certificate | 14 | 16 | 11 | 13 | 12 | 16 | 6.8 | 10.1 |
| Postgraduate | 22 | 29 | 21 | 24 | 23 | 19 | 11.5 | 8.5 |
| Other | 1 | 1 | 0 | 2 | 1 | 0 | 0.4 | - |
| **Country of Birth** |  |  |  |  |  |  |  |  |
| Australia | 151 | 152 | 156 | 167 | 160 | 162 | 78.7 | 65.6 |
| Overseas | 50 | 48 | 45 | 34 | 40 | 39 | 28.3 | 34.4 |
| **Gender** |  |  |  |  |  |  |  |  |
| Woman | 103 | 99 | 94 | 102 | 106 | 104 | 50.5 | 50.7 |
| Man | 98 | 101 | 105 | 99 | 94 | 97 | 49.3 | 49.3 |
| Non-binary / other | 0 | 0 | 2 | 0 | 0 | 0 | 0.2 | - |
| **Age bracket (years)** |  |  |  |  |  |  |  |  |
| 18-24 | 20 | 24 | 20 | 22 | 21 | 19 | 10.5 | 12.1 |
| 25-34 | 33 | 39 | 51 | 42 | 37 | 35 | 19.7 | 20.6 |
| 35-44 | 39 | 41 | 27 | 38 | 33 | 32 | 17.4 | 19.3 |
| 45-54 | 36 | 29 | 30 | 33 | 37 | 37 | 16.8 | 18.0 |
| 55-64 | 28 | 29 | 31 | 25 | 36 | 30 | 14.9 | 16.6 |
| 65+ | 45 | 38 | 42 | 41 | 36 | 48 | 20.8 | 13.3 |
| **Vaccine / COVID-19 History** |  |  |  |  |  |  |  |  |
| **Previous COVID-19 Vaccine** |  |  |  |  |  |  |  |  |
| AstraZeneca (both doses) | 84 | 84 | 84 | 84 | 83 | 84 | 41.8 | - |
| Pfizer (both doses) | 116 | 115 | 116 | 116 | 116 | 115 | 57.6 | - |
| AstraZeneca and<br>Pfizer (combination) | 1 | 1 | 1 | 1 | 1 | 2 | 06 | - |
| **Months since last COVID-19 vaccination** |  |  |  |  |  |  |  |  |
| 0-5 | 188 | 185 | 183 | 176 | 185 | 180 | 91.1 | - |
| 6-15 | 12 | 13 | 15 | 22 | 12 | 19 | 7.7 | - |
| 16-20 | 1 | 2 | 3 | 3 | 3 | 2 | 1.2 | - |
| **COVID-19 Exposure: Personal infection** |  |  |  |  |  |  |  |  |
| Yes | 7 | 4 | 5 | 2 | 6 | 3 | 2.2 | - |
| No | 194 | 196 | 196 | 199 | 194 | 198 | 97.8 | - |
| **COVID-19 Exposure: Significant others** |  |  |  |  |  |  |  |  |
| Yes | 28 | 24 | 24 | 25 | 21 | 24 | 12.1 | - |
| No | 173 | 176 | 177 | 176 | 197 | 177 | 89.4 | - |
| **Previously heard of the Pfizer COVID-19 vaccine** |  |  |  |  |  |  |  |  |
| Yes | 198 | 198 | 198 | 196 | 198 | 200 | 98.7 | - |
| No | 3 | 2 | 3 | 5 | 2 | 1 | 1.3 | - |
| **Previous heard of the Moderna COVID-19 vaccine** |  |  |  |  |  |  |  |  |
| Yes | 183 | 185 | 181 | 178 | 174 | 187 | 90.4 | - |
| No | 18 | 15 | 20 | 23 | 26 | 14 | 9.6 | - |

**Figure 3:**
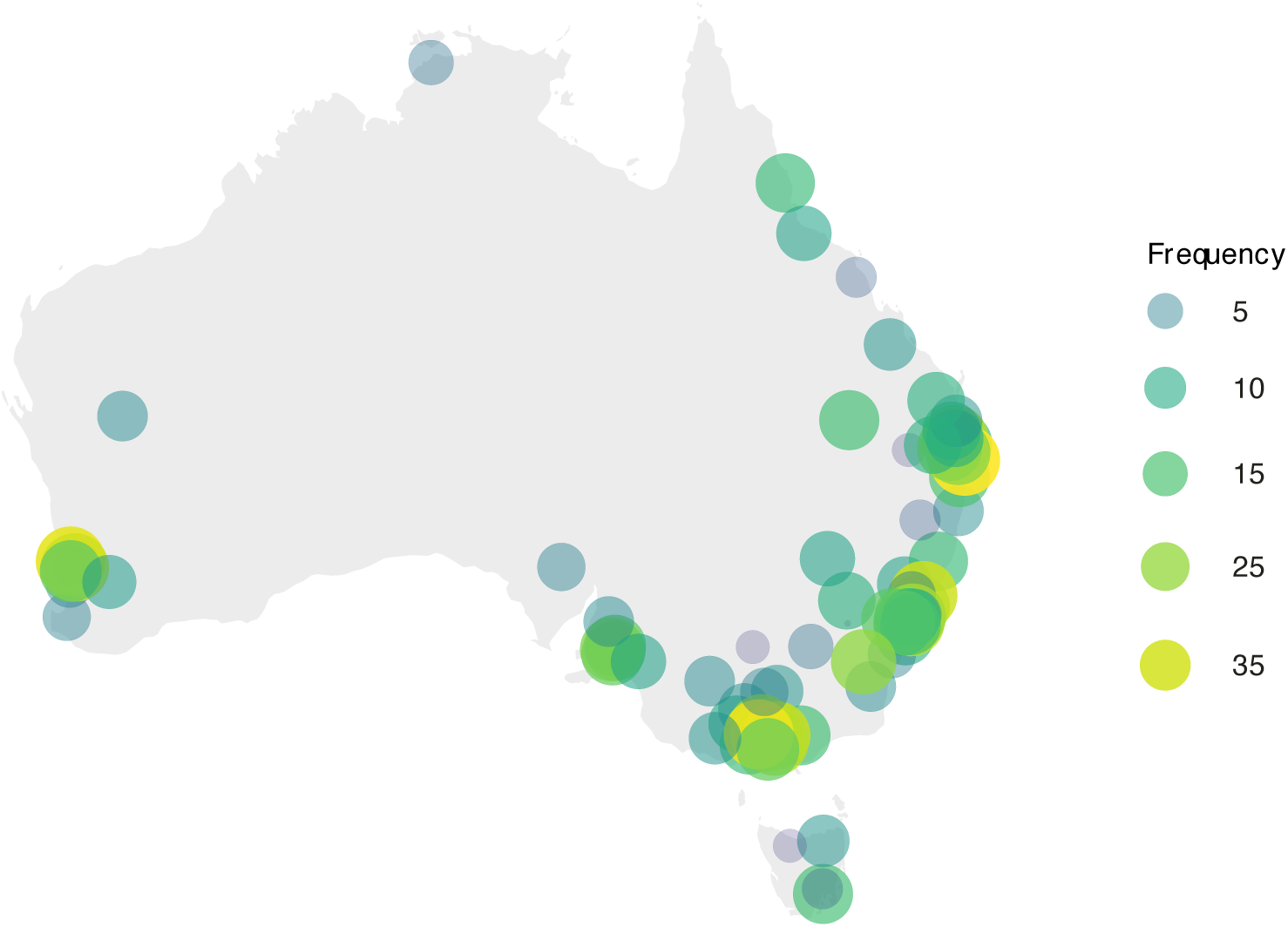
Geospatial data with participant frequency plotted against SA4 postal regions in Australia

#### Manipulation Checks

As expected, side effect familiarity was greater for the Pfizer than Moderna vaccine (*t*(1203)=28.63, *p*<.0001, Cohen’s *d*_*z*_ =.83). During the intervention, more participants underestimated side effects for the Modena (*N*=118) relative to Pfizer (*N*=65) vaccine (*χ*^*2*^(1) = 18.97, *p*<.0001), due to the vaccine having a higher incidence of side effects than Pfizer.

#### Primary Analysis

Booster Intentions were higher for any Framing relative to Control (Contrast1: *F*(1, 1192)=11.56, *p*=.0007, *η*_*p*_^*2*^=.010) and for Positive Framing (*M*=75.7, *SE*=0.9, 95% CIs[73.9, 77.4]) relative to Negative Framing (*M*=70.7, *SE*=0.9, 95% CIs[68.9, 72.4]; *F*(1, 1192)=4.68, *p*=.031, *η*_*p*_^*2*^=.004). As presented in Figure 4a, the anticipated Framing*Vaccine interaction was not significant at Contrast2 (Positive vs. Negative: *F*(1, 1192)=3.31, *p*=.069, *η*_*p*_^*2*^=.003), but was at Contrast1 (Framing vs. Control: *F*(1, 1192)=8.91, *p*=.003, *η*_*p*_^*2*^=.007), with the effect of any information over Control larger for the Unfamiliar vaccine (see S1.6 for full model output and means).

**Figure 4:**
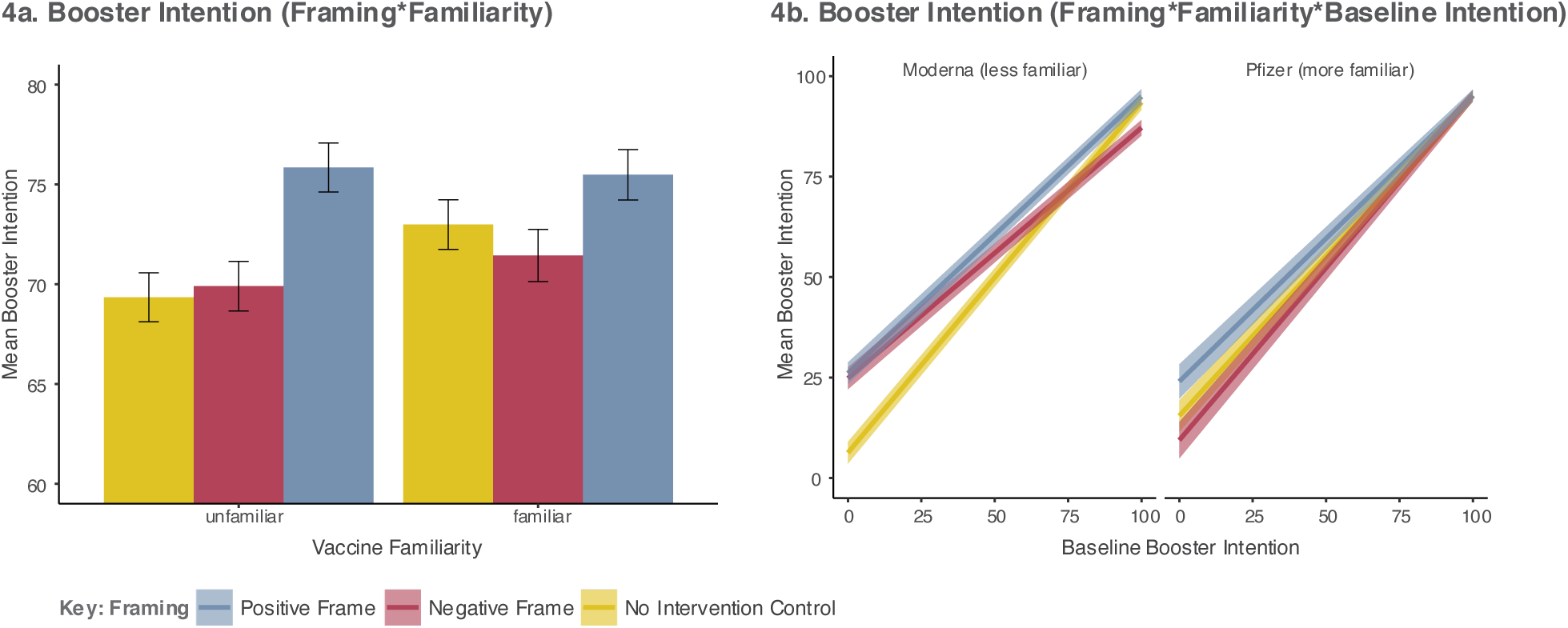
Model estimated mean differences in the primary outcome (Booster Intention). 4a depicts mean Booster Intention for each framing condition by vaccine type. 4b depicts the Framing*Familiarity*Baseline Booster Intention interaction. All error bars represent ± 1SEM.

As in Figure 4b, a three-way Framing*Vaccine*Baseline Intention interaction was present at both Contrasts (Control vs. Framing: *F*(1, 1192)=7.39, *p*=.007, *η*_*p*_^*2*^=.006 | Positive vs. Negative: *F*(2, 1192)=5.26, *p*=.022, *η*_*p*_^*2*^=.004). For the less familiar Moderna vaccine, Positive Framing (*M*=94.92, *SE*=1.88, 95% CI[91.24, 98.60]) increased Booster Intention relative to Negative Framing (*M*=87.18, *SE*=1.95, 95% CI[83.35, 91.0]) at high Baseline Intent (VAS=100). For the more familiar Pfizer vaccine, Positive Framing (*M*=24.03, *SE*=4.29, 95% CI[15.62, 32.4]) increased Intention at low Baseline Intent (VAS=0) relative to Negative Framing (*M*=9.42, *SE*=4.54, 95% CI[0.51, 18.3]). Results appeared not to be driven by greater underestimation for the less familiar Moderna vaccine (see: S1.7).

#### Secondary-Predictors: Mediation

Positive Framing is theorised to create a valence-consistent shift in perceptions that alters evaluation and intention^16,17^. Analysis was therefore run to explore whether side effect perceptions mediated the effect of framing (Positive vs. Negative) on Booster Intentions across vaccines (i.e., as if the intervention were generally applied). Three models were run with baseline-corrected Booster Intention (change score) as the outcome and baseline-corrected secondary predictors as mediators. As in Table 2, partial mediation was observed through a decrease in Side Effect Worry and Severity associated with Positive Framing. This was specific to side effect perceptions and not significant for Booster Protection.

**Table 2:** Mediation of Framing (Positive vs. Negative) on Booster Intention through Side Effect Worry, Perception of Side Effect Severity, and Perceived Booster Protection. Paths a and b represent paths from predictor to mediator and mediator to outcome. Path c represents the total effect (Framing on Booster Intention), c’ the association between Framing and Booster Intention controlling for all other paths, and a*b indirect effect of the mediator on the Framing – Booster Intention association. Bias-corrected bootstrapped 95% CIs (10,000 samples) are presented.

| | $\beta$ | *B* | *SE B* | *Z* | *p* | 95% CIs |
| --- | --- | --- | --- | --- | --- | --- |
| a | -0.14 | -6.51 | 1.66 | -3.92 | **<.001** | [-9.85, -3.36] |
| b | -0.18 | -0.15 | 0.04 | -3.77 | **<.001** | [-0.18, -0.18] |
| c | 0.10 | 3.93 | 1.37 | 2.87 | **.004** | [1.22, 6.55] |
| c' | 0.08 | 2.98 | 1.34 | 2.23 | **.026** | [0.33, 5.56] |
| a*b | 0.02 | 0.95 | 0.35 | 2.68 | **.007** | [0.40, 1.82] |

B) Mediation Model: Perceived Side Effect Severity
| | $\beta$ | *B* | *SE B* | *Z* | *p* | 95% CIs |
| --- | --- | --- | --- | --- | --- | --- |
| a | -0.20 | -8.63 | 1.48 | -5.83 | **<.001** | [-11.54, -5.66] |
| b | -0.20 | -0.18 | 0.05 | -3.67 | **<.001** | [-0.29, -0.09] |
| c | 0.10 | 3.93 | 1.37 | 2.87 | **.004** | [1.22, 6.55] |
| c' | 0.06 | 2.34 | 1.37 | 1.71 | .088 | [-0.36, 4.99] |
| a*b | 0.04 | 1.59 | 0.52 | 3.02 | **.002** | [0.04, 0.08] |

C) Mediation Model: Perceived Booster Protection
| | $\beta$ | *B* | *SE B* | *Z* | *p* | 95% CIs |
| --- | --- | --- | --- | --- | --- | --- |
| a | 0.05 | 1.74 | 1.16 | 1.50 | .134 | [-0.53, 3.99] |
| b | 0.32 | 0.38 | 0.06 | 5.88 | **<.001** | [0.25, 0.50] |
| c | 0.10 | 3.93 | 1.37 | 1.47 | **.004** | [1.22, 6.55] |
| c' | 0.09 | 3.28 | 1.30 | 2.52 | **.012** | [0.70, 5.75] |
| a*b | 0.02 | 0.66 | 0.45 | 1.47 | .143 | [-0.15, 1.62] |

## Discussion

The present study tested an intervention involving the presentation of framed side-effect information. Overall, providing side effect information of any type to participants increased their COVID-19 Booster Intention relative to the Control group, who received no information at all. Of note, Positive Framing further increased Booster Intention relative to Negative Framing by 5 percentage points. While the mapping between intention and uptake is unlikely to be exact^18^ (see below), our results suggest that Positive Framing could lead to up to half a million extra booster vaccinations among those aged 18 and over (based on population averages from the ABS Education and Work Dataset 2021). Results therefore indicate that presenting positively-framed information to people, for example on Australian Government and AusVaxSafety websites, is likely to increase favorable perceptions of booster vaccination.

Based on our previous research^6^ and the broader literature concerning framing effects on vaccine-related intentions^7,8^, vaccine familiarity was anticipated to modulate the effect of Positive Framing. However, Booster Intention after Positive Framing was only numerically, not statistically, larger for the less familiar Moderna vaccine compared to Negative Framing. Instead, we found that both Positive and Negative provided an increased benefit to Booster Intention over Control for the less familiar Moderna vaccine relative to the more familiar Pfizer vaccine (although this appeared to be driven by the Positive Framing manipulation). A three-way interaction involving Baseline Intention qualified these effects. Here, the difference between Positive and Negative Framing was largest at low levels of Baseline Intent for the Familiar vaccine, but at high levels of Baseline Intent for the Unfamiliar vaccine. In our previous research, Positive Framing was found to decrease Booster Intention relative to Negative Framing for familiar vaccines^6^. Notably, this was not the case here. Several differences exist between these studies, such as sample location, number of framed side effects, and the mode of presentation (PILs vs. infographics). As such, future research should strive to understand the conditions under which Positive Framing reduces vaccine intention to ensure that optimal messaging is employed.

Population level public health information on side effects is inherently general and cannot contain nuanced information about the influence of prior history of vaccination or hesitancy. While a complex pattern of results was observed, Positive Framing was always either superior, or equal to, Negative Framing and Control at all levels of Baseline Intent for both vaccines. Moreover, at differing levels of Baseline Intent, Negative Framing decreased Booster Intention relative to Positive Framing for both familiar and less familiar vaccines. Among those most resistant to vaccination, Positive Framing of the more familiar vaccine (Pfizer) increased Booster Intention by 14.6 percentage-points relative to Negative Framing, and by 19.8 percentage-points relative to Control for the less familiar vaccine (Moderna). As such, there was never any disadvantage to employing Positive Framing in the present sample, even when targeting those with low Baseline Intent; a population where increasing vaccine acceptance is of critical importance^19^. This suggests that Positive Framing is the optimal presentation mode.

In terms of mechanisms, theories of attribute framing posit that Positive Framing results in a valence-consistent shift in perception that alters evaluation^16,17^. We therefore reasoned that Positive Framing would alter side effect perception, increasing intention to be vaccinated. Mediation analysis provided tentative support for this theory. Specifically, a reduction in the perception of side effect severity and side effect worry partially mediated the effect of Positive (vs. Negative) Framing on Booster Intention, suggesting that Framing may indeed reduce side effect hesitancy. Mediation was not found through perceived booster protection, with the association between Framing and booster protection weaker than with side effect perceptions (severity and worry).

There are several strengths to the present study, including the framing of real COVID-19 vaccines and the use of actual side effect data presented to the Australian population. Especially as much of the literature in this area frames fictitious vaccines and asks participants to imagine scenarios that they have never experienced and may be unlikely to ever experience^7,8,12,17,20^. Limitations of the study include the measurement of intention, but not uptake. While intention has been found to predict vaccination^21-24^, longitudinal research is needed to directly assess the role of framing on actual uptake, as well as the longevity of the framing effect among those yet to receive a booster vaccine. Relevant to the lag in booster uptake in Australia^25^, the present research focused on increasing intention among those already receiving a primary course of COVID-19 vaccination. However, these results do not speak to the effect of framing on those never vaccinated. Investigation of framing on vaccine intention at all points of the vaccination programme would provide a more comprehensive account of the effect of framing on vaccine intentions in general.

In summary, a brief online intervention engaging participants in side effect estimation before presenting positively framed side effect information can increase booster vaccine intention. Given the ease with which Positive Framing can be implemented, combined with the fact that the presentation of statistical information in this format does not violate patient informed consent^9^, the potential exists for framing of this type to make a real difference in improving societal protection from COVID-19 through reduced vaccine hesitancy.

## Data Availability

All data produced are available online at https://osf.io/nfxr3/?view_only=e0717d3f063245268f8ce88144afaa3f

## Data Availability

The code and raw data necessary to replicate the reported analysis is available through the Open Science Framework repository: https://osf.io/nfxr3/?view_only=e0717d3f063245268f8ce88144afaa3f

## Author contributions

KB, BC and KF conceived the experimental design. KB was responsible for collecting and analysing the data. KB wrote the first draft of the article. BC and KF edited and contributed to the final version.

