## Supplementary Materials for "The impact of side effect framing on COVID-19 booster vaccine intentions in an Australian sample"

#### Contents

**S1.1: Analysis of Secondary Predictors** 2

**S1.2: STROBE Checklist** 14

**S1.3: Survey Structure** 16

**S1.4: Infographics used in the study** 26

**S1.5: Pre-analysis data cleaning** 34

**S1.6: Primary Analysis - Full Statistical Model** 35

**S1.7: Supplementary Model Removing Underestimators** 37

### S1.1: Analysis of Secondary Predictors

Side effect worry, perceived side effect severity, and perceived booster vaccine protection associated with the framed vaccine were measured pre- and post-manipulation. Secondary outcomes were measured to test for mediation. However, pre-registered ANCOVA models (consistent with the primary outcome) were first run to explore whether similar patterns in the data existed.

#### S1.1.1: Side Effect Worry (ANCOVA)

##### Summary

A 3(Framing) * 2(Familiarity) ANCOVA, including interactions with the covariate (Baseline Side Effect Worry), revealed a main effect of Framing ($F(2, 1192)=12.14$, $p<.00001$, *np2*=.02), present in Contrast 1 (Control vs. Framing:$F(1, 1192)=10.97$, $p=.001$, *np2*=.01) and Contrast 2 (Positive vs. Negative:$F(1, 1192)=13.67$, $p=.0002$, *np2*=.01). As depicted in Figure 1.1.1 below, there was a reduction in side effect worry associated with the Positive Frame.

The highest order interaction involved Framing, Familiarity, and Baseline Side Effect Worry ($F(2, 1192)=5.85$, $p=.003$, *np2*=.01), present at Contrast 1 (Control vs. Framing:$F(1, 1192)=5.56$, $p=.019$, *np2*=.005) and Contrast 2 (Positive vs. Negative:$F(1, 1192)=6.75$, $p=.01$, *np2*=.006), as depicted in Figure 1.1.2 below.

##### Item wording:

"How worried would you be about experiencing side effects after receiving [framed vaccine]"

##### Variable names:

- seworry_t2 (side effect worry associated with the framed vaccine post-intervention)
- seworry_fv_t1 (side effect worry associated with the framed vaccine at baseline)

#### Overall effects

| | Sum Sq | Df | F value | Pr(>F) |
|---|---|---|---|---|
| (Intercept) | 48322.332 | 1 | 127.2427 | 0.0000 |
| seworry_fv_t1 | 455609.140 | 1 | 1199.7127 | 0.0000 |
| frame | 9224.114 | 2 | 12.1445 | 0.0000 |
| fam | 3436.624 | 1 | 9.0493 | 0.0027 |
| seworry_fv_t1:frame | 7773.289 | 2 | 10.2343 | 0.0000 |
| seworry_fv_t1:fam | 1527.976 | 1 | 4.0235 | 0.0451 |
| frame:fam | 5311.089 | 2 | 6.9926 | 0.0010 |
| seworry_fv_t1:frame:fam | 4443.141 | 2 | 5.8499 | 0.0030 |
| Residuals | 452680.109 | 1192 | NA | NA |

| | eta.sq | eta.sq.part |
|---|---|---|
| seworry_fv_t1 | 0.4752 | 0.5016 |
| frame | 0.0096 | 0.0200 |
| fam | 0.0036 | 0.0075 |
| seworry_fv_t1:frame | 0.0081 | 0.0169 |
| seworry_fv_t1:fam | 0.0016 | 0.0034 |
| frame:fam | 0.0055 | 0.0116 |
| seworry_fv_t1:frame:fam | 0.0046 | 0.0097 |

#### Contrasts

| | Sum Sq | Df | F value | Pr(>F) |
|---|---|---|---|---|
| (Intercept) | 48322.3317 | 1 | 127.2427 | 0.0000 |
| seworry_fv_t1 | 455609.1399 | 1 | 1199.7127 | 0.0000 |
| fam | 3436.6239 | 1 | 9.0493 | 0.0027 |
| cont1 | 4167.6357 | 1 | 10.9742 | 0.0010 |
| cont2 | 5189.7441 | 1 | 13.6657 | 0.0002 |
| seworry_fv_t1:fam | 1527.9762 | 1 | 4.0235 | 0.0451 |
| seworry_fv_t1:cont1 | 7142.8585 | 1 | 18.8086 | 0.0000 |
| fam:cont1 | 2948.7318 | 1 | 7.7646 | 0.0054 |
| seworry_fv_t1:cont2 | 869.3684 | 1 | 2.2892 | 0.1305 |
| fam:cont2 | 2439.2090 | 1 | 6.4229 | 0.0114 |
| seworry_fv_t1:fam:cont1 | 2112.0330 | 1 | 5.5614 | 0.0185 |
| seworry_fv_t1:fam:cont2 | 2561.8028 | 1 | 6.7458 | 0.0095 |
| Residuals | 452680.1087 | 1192 | NA | NA |

| | eta.sq | eta.sq.part |
|---|---|---|
| seworry_fv_t1 | 0.4752 | 0.5016 |
| fam | 0.0036 | 0.0075 |
| cont1 | 0.0043 | 0.0091 |
| cont2 | 0.0054 | 0.0113 |
| seworry_fv_t1:fam | 0.0016 | 0.0034 |
| seworry_fv_t1:cont1 | 0.0074 | 0.0155 |
| fam:cont1 | 0.0031 | 0.0065 |
| seworry_fv_t1:cont2 | 0.0009 | 0.0019 |
| fam:cont2 | 0.0025 | 0.0054 |
| seworry_fv_t1:fam:cont1 | 0.0022 | 0.0046 |
| seworry_fv_t1:fam:cont2 | 0.0027 | 0.0056 |

#### Group Means

| frame | emmean | SE | df | lower.CL | upper.CL |
|---|---|---|---|---|---|
| control | 38.87639 | 0.97434 | 1192 | 36.96477 | 40.78801 |
| negative | 41.46366 | 0.97672 | 1192 | 39.54738 | 43.37994 |
| positive | 35.56721 | 0.97408 | 1192 | 33.65611 | 37.47831 |

| fam | emmean | SE | df | lower.CL | upper.CL |
|---|---|---|---|---|---|
| unfamiliar | 39.97630 | 0.79550 | 1192 | 38.41557 | 41.53703 |
| familiar | 37.29521 | 0.79675 | 1192 | 35.73203 | 38.85840 |

| frame | fam | emmean | SE | df | lower.CL | upper.CL |
|---|---|---|---|---|---|---|
| control | unfamiliar | 38.99626 | 1.381285 | 1192 | 36.28624 | 41.70628 |
| negative | unfamiliar | 43.88088 | 1.375167 | 1192 | 41.18286 | 46.57889 |
| positive | unfamiliar | 37.05175 | 1.377069 | 1192 | 34.35000 | 39.75350 |
| control | familiar | 38.75652 | 1.374565 | 1192 | 36.05968 | 41.45336 |
| negative | familiar | 39.04644 | 1.387390 | 1192 | 36.32444 | 41.76844 |
| positive | familiar | 34.08267 | 1.378043 | 1192 | 31.37901 | 36.78633 |

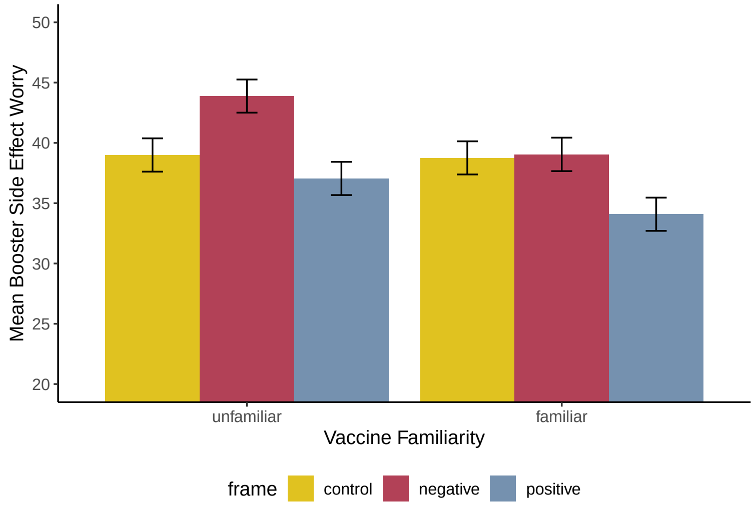


Figure 1.1.1: Graph depicting means for the Framing * Familiarity Interaction (error bars represented +/- 1 SEM)

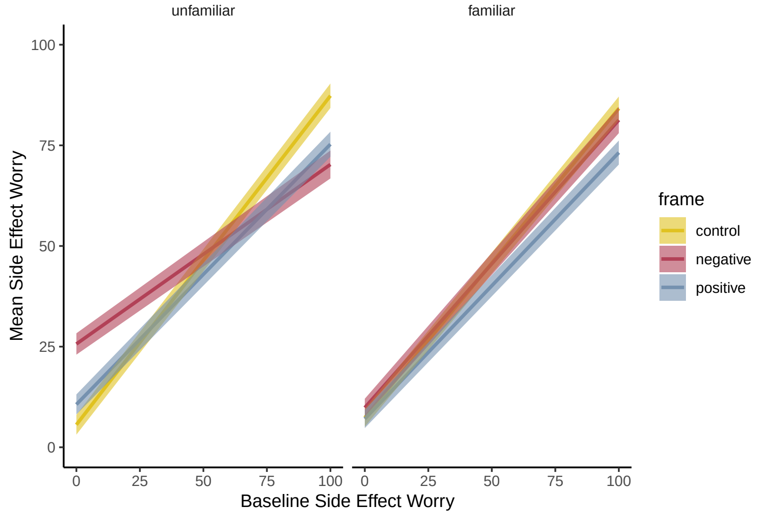


Figure 1.1.2: Graph depicting the Framing * Familiarity * Baseline Side Effect Worry Interaction

#### S1.1.2: Secondary Predictors - Side Effect Severity (ANCOVA)

##### Summary

A 3(Framing) * 2(Familiarity) ANCOVA, including interactions with the covariate (Baseline Side Effect Severity), revealed a main effect of Framing ($F(2, 1192)=11.08$, $p=.00002$, *np2*=.02), present in Contrast 1 (Control vs. Framing:$F(1, 1192)=8.77$, $p=.003$, *np2*=.01) and Contrast 2 (Positive vs. Negative:$F(1, 1192)=13.55$, $p=.0002$, *np2*=.01). As depicted in Figure 1.2.1 below, there was a reduction in perceived side effect severity associated with the Positive Frame.

Higher order Framing * Familiarity and Framing * Familiarity * Baseline Side Effect Severity interactions did not reach statistical significance.

##### Item wording:

"Overall, how severe do you think the [framed vaccine] side effects are?

##### Variable names:

- sesev_t2 (perceived side effect severity associated with the framed vaccine post-intervention)
- sesev_fv_t1 (perceived side effect severity associated with the framed vaccine at baseline)

#### Overall effects

| | Sum Sq | Df | F value | Pr(>F) |
|---|---|---|---|---|
| (Intercept) | 39133.730 | 1 | 129.02452 | 0.00000 |
| sesev_fv_t1 | 318733.957 | 1 | 1050.87088 | 0.00000 |
| frame | 6719.070 | 2 | 11.07644 | 0.00002 |
| fam | 4688.379 | 1 | 15.45766 | 0.00009 |
| sesev_fv_t1:frame | 9001.559 | 2 | 14.83914 | 0.00000 |
| sesev_fv_t1:fam | 2923.839 | 1 | 9.63994 | 0.00195 |
| frame:fam | 1231.300 | 2 | 2.02981 | 0.13181 |
| sesev_fv_t1:frame:fam | 1038.915 | 2 | 1.71266 | 0.18083 |
| Residuals | 361539.066 | 1192 | NA | NA |

| | eta.sq | eta.sq.part |
|---|---|---|
| sesev_fv_t1 | 0.43833 | 0.46854 |
| frame | 0.00924 | 0.01825 |
| fam | 0.00645 | 0.01280 |
| sesev_fv_t1:frame | 0.01238 | 0.02429 |
| sesev_fv_t1:fam | 0.00402 | 0.00802 |
| frame:fam | 0.00169 | 0.00339 |
| sesev_fv_t1:frame:fam | 0.00143 | 0.00287 |

#### Contrasts

| | Sum Sq | Df | F value | Pr(>F) |
|---|---|---|---|---|
| (Intercept) | 39133.72958 | 1 | 129.02452 | 0.00000 |
| sesev_fv_t1 | 318733.95744 | 1 | 1050.87088 | 0.00000 |
| fam | 4688.37905 | 1 | 15.45766 | 0.00009 |
| cont1 | 2660.17034 | 1 | 8.77062 | 0.00312 |
| cont2 | 4109.97569 | 1 | 13.55065 | 0.00024 |
| sesev_fv_t1:fam | 2923.83918 | 1 | 9.63994 | 0.00195 |
| sesev_fv_t1:cont1 | 8999.75800 | 1 | 29.67234 | 0.00000 |
| fam:cont1 | 1229.48179 | 1 | 4.05362 | 0.04430 |
| sesev_fv_t1:cont2 | 45.55970 | 1 | 0.15021 | 0.69840 |
| fam:cont2 | 2.62932 | 1 | 0.00867 | 0.92583 |
| sesev_fv_t1:fam:cont1 | 1019.54028 | 1 | 3.36144 | 0.06699 |
| sesev_fv_t1:fam:cont2 | 6.62358 | 1 | 0.02184 | 0.88254 |
| Residuals | 361539.06577 | 1192 | NA | NA |

| | eta.sq | eta.sq.part |
|---|---|---|
| sesev_fv_t1 | 0.43833 | 0.46854 |
| fam | 0.00645 | 0.01280 |
| cont1 | 0.00366 | 0.00730 |
| cont2 | 0.00565 | 0.01124 |
| sesev_fv_t1:fam | 0.00402 | 0.00802 |
| sesev_fv_t1:cont1 | 0.01238 | 0.02429 |
| fam:cont1 | 0.00169 | 0.00339 |
| sesev_fv_t1:cont2 | 0.00006 | 0.00013 |
| fam:cont2 | 0.00000 | 0.00001 |
| sesev_fv_t1:fam:cont1 | 0.00140 | 0.00281 |
| sesev_fv_t1:fam:cont2 | 0.00001 | 0.00002 |

#### Group Means

| frame | emmean | SE | df | lower.CL | upper.CL |
|---|---|---|---|---|---|
| control | 36.57414 | 0.87006 | 1192 | 34.86712 | 38.28115 |
| negative | 37.36624 | 0.87187 | 1192 | 35.65567 | 39.07682 |
| positive | 29.90409 | 0.87104 | 1192 | 28.19515 | 31.61304 |

| fam | emmean | SE | df | lower.CL | upper.CL |
|---|---|---|---|---|---|
| unfamiliar | 35.83656 | 0.71043 | 1192 | 34.44274 | 37.23039 |
| familiar | 33.39309 | 0.71190 | 1192 | 31.99638 | 34.78979 |

| frame | fam | emmean | SE | df | lower.CL | upper.CL |
|---|---|---|---|---|---|---|
| control | unfamiliar | 37.16098 | 1.232356 | 1192 | 34.74315 | 39.57880 |
| negative | unfamiliar | 39.14614 | 1.228493 | 1192 | 36.73589 | 41.55639 |
| positive | unfamiliar | 31.20257 | 1.230634 | 1192 | 28.78812 | 33.61702 |
| control | familiar | 35.98730 | 1.228542 | 1192 | 33.57695 | 38.39764 |
| negative | familiar | 35.58635 | 1.237518 | 1192 | 33.15839 | 38.01430 |
| positive | familiar | 28.60562 | 1.233040 | 1192 | 26.18645 | 31.02479 |

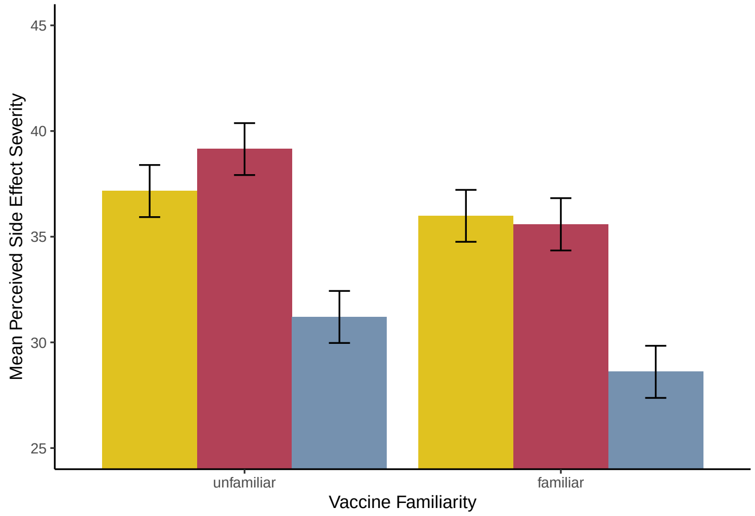


Figure 1.2.1: Graph depicting means for the Framing * Familiarity Interaction (error bars represented +/- 1 SEM)

#### S1.1.3: Secondary Predictors - Perceived protection (ANCOVA)

##### Summary

There was no main effect of Framing ($F$(2, 1192)=2.24, $p$=.107, *np2*=.004), but an effect of Framing at Contrast 2 (Positive vs. Negative:$F$(1, 1192)=3.93, $p$=.048, *np2*=.003), which was driven by Positive Framing increasing the perception of protection for the Unfamiliar vaccine. As depicted in Figure 1.3.1 below, there was a Framing * Familiarity interaction ($F$(2, 1192)=6.14, $p$=.002, *np2*=.01), but this was only present at Contrast 1 (Control vs. Framing:$F$(1, 1192)=11.12, $p$=.0009, *np2*=.01).

The highest order interaction involved Framing, Familiarity, and Baseline Side Effect Worry ($F$(2, 1192)=6.66, $p$=.001, *np2*=.01). Again, this was present at Contrast 1 only (Control vs. Framing:$F$(1, 1192)=11.00, $p$=.0009, *np2*=.01), see Figure 1.3.2 below.

##### Item wording:

"How confident would you feel that you had all the protection you need after receiving the [framed] booster?"

##### Variable names:

- protect_t2 (perceived protection associated with the framed vaccine post-intervention)
- protect_fv_t1 (perceived protection associated with the framed vaccine at baseline)

#### Overall effects

| | Sum Sq | Df | F value | Pr(>F) |
|---|---|---|---|---|
| (Intercept) | 39864.88409 | 1 | 164.90522 | 0.00000 |
| protect_fv_t1 | 484630.33629 | 1 | 2004.72350 | 0.00000 |
| frame | 1081.33605 | 2 | 2.23653 | 0.10728 |
| fam | 42.66857 | 1 | 0.17650 | 0.67447 |
| protect_fv_t1:frame | 1893.91144 | 2 | 3.91718 | 0.02015 |
| protect_fv_t1:fam | 192.29187 | 1 | 0.79544 | 0.37264 |
| frame:fam | 2966.32998 | 2 | 6.13527 | 0.00223 |
| protect_fv_t1:frame:fam | 3220.44585 | 2 | 6.66085 | 0.00133 |
| Residuals | 288159.12047 | 1192 | NA | NA |

| | eta.sq | eta.sq.part |
|---|---|---|
| protect_fv_t1 | 0.60506 | 0.62712 |
| frame | 0.00135 | 0.00374 |
| fam | 0.00005 | 0.00015 |
| protect_fv_t1:frame | 0.00236 | 0.00653 |
| protect_fv_t1:fam | 0.00024 | 0.00067 |
| frame:fam | 0.00370 | 0.01019 |
| protect_fv_t1:frame:fam | 0.00402 | 0.01105 |

#### Contrasts

| | Sum Sq | Df | F value | Pr(>F) |
|---|---|---|---|---|
| (Intercept) | 39864.8841 | 1 | 164.9052 | 0.0000 |
| protect_fv_t1 | 484630.3363 | 1 | 2004.7235 | 0.0000 |
| fam | 42.6686 | 1 | 0.1765 | 0.6745 |
| cont1 | 147.8757 | 1 | 0.6117 | 0.4343 |
| cont2 | 949.8237 | 1 | 3.9290 | 0.0477 |
| protect_fv_t1:fam | 192.2919 | 1 | 0.7954 | 0.3726 |
| protect_fv_t1:cont1 | 57.1577 | 1 | 0.2364 | 0.6269 |
| fam:cont1 | 2689.3713 | 1 | 11.1249 | 0.0009 |
| protect_fv_t1:cont2 | 1853.6280 | 1 | 7.6677 | 0.0057 |
| fam:cont2 | 317.1274 | 1 | 1.3118 | 0.2523 |
| protect_fv_t1:fam:cont1 | 2654.2232 | 1 | 10.9795 | 0.0009 |
| protect_fv_t1:fam:cont2 | 637.1026 | 1 | 2.6354 | 0.1048 |
| Residuals | 288159.1205 | 1192 | NA | NA |

| | eta.sq | eta.sq.part |
|---|---|---|
| protect_fv_t1 | 0.6051 | 0.6271 |
| fam | 0.0001 | 0.0001 |
| cont1 | 0.0002 | 0.0005 |
| cont2 | 0.0012 | 0.0033 |
| protect_fv_t1:fam | 0.0002 | 0.0007 |
| protect_fv_t1:cont1 | 0.0001 | 0.0002 |
| fam:cont1 | 0.0034 | 0.0092 |
| protect_fv_t1:cont2 | 0.0023 | 0.0064 |
| fam:cont2 | 0.0004 | 0.0011 |
| protect_fv_t1:fam:cont1 | 0.0033 | 0.0091 |
| protect_fv_t1:fam:cont2 | 0.0008 | 0.0022 |

#### Group Means

| protect_fv_t1 | emmean | SE | df | lower.CL | upper.CL |
|---|---|---|---|---|---|
| 68.74917 | 70.23068 | 0.45179 | 1192 | 69.34429 | 71.11706 |

| frame | emmean | SE | df | lower.CL | upper.CL |
|---|---|---|---|---|---|
| control | 69.64774 | 0.78603 | 1192 | 68.10558 | 71.18991 |
| negative | 69.57160 | 0.77978 | 1192 | 68.04171 | 71.10149 |
| positive | 71.47268 | 0.78174 | 1192 | 69.93895 | 73.00641 |

| fam | emmean | SE | df | lower.CL | upper.CL |
|---|---|---|---|---|---|
| unfamiliar | 69.69850 | 0.63931 | 1192 | 68.44420 | 70.95279 |
| familiar | 70.76286 | 0.63854 | 1192 | 69.51007 | 72.01564 |

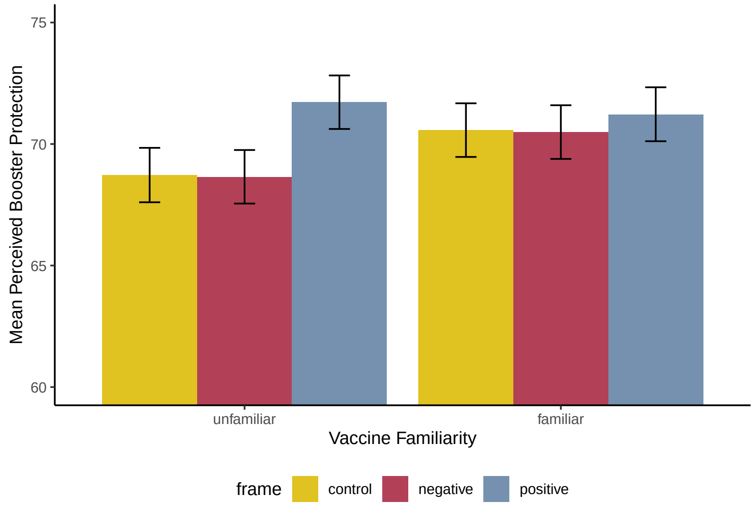


Figure 1.3.1: Graph depicting means for the Framing * Familiarity Interaction (error bars represented +/- 1 SEM)

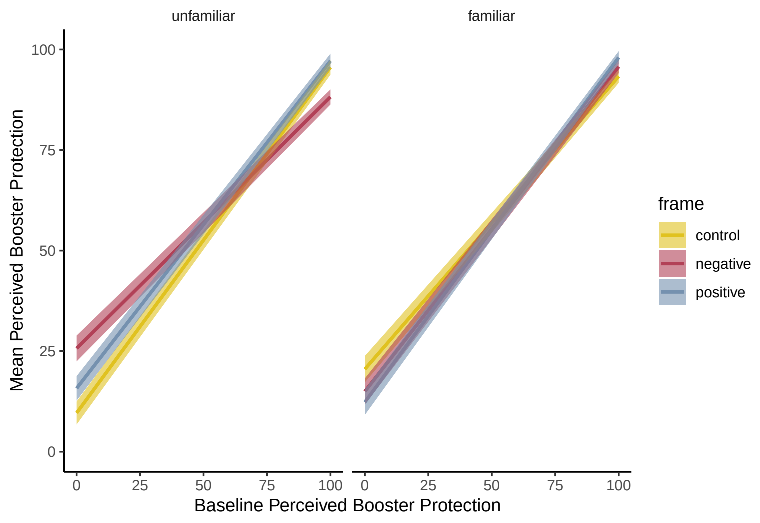


Figure 1.3.2: Graph depicting the Framing * Familiarity * Baseline Protection Interaction

#### S1.1.4: Generalisation of the Framing Effect

**Item wording:**

"If you were offered the [vaccine type not framed] as a booster how likely would you be to accept it?

**Variable names:**

- intent_t2gen (intention to receive the booster vaccine not framed post-intervention)
- intent_fv_t1gen (intention to receive the booster vaccine not framed at baseline)

**Summary**

Overall, there was a main effect of Framing ($F$(2, 1192)=3.28, $p$=.038, *np2*=.005), present in Contrast 1 only (Control vs. Framing: $F$(1, 1192)=6.55, $p$=.011, *np2*=.005) and a Framing * Familiarity interaction ($F$(2, 1192)=5.41, $p$=.005, *np2*=.009), present in Contrast 1 only (Control vs. Framing: $F$(1, 1192)=10.70, $p$=.001, *np2*=.009).

The highest order interaction was a Framing * Familiarity * Baseline Intention interaction ($F$(2, 1192)=5.81, $p$=.003, *np2*=.010) and was also present in Contrast 1 only (Control vs. Framing: $F$(1, 1192)=11.60, $p$=.0007, *np2*=.010). As depicted in Figure 1.4.2 below, there was an effect of Framing, relative to Control, at low Baseline Intention (for the non-framed vaccine), but only among those who received framing for the Unfamiliar vaccine (i.e., the effect of framing generalised from the Unfamiliar to the Familiar vaccine, but not vice versa).

#### Overall effects

| | Sum Sq | Df | F value | Pr(>F) |
|---|---|---|---|---|
| (Intercept) | 19226.0356 | 1 | 86.91001 | 0.00000 |
| intent_fv_t1gen | 537629.8066 | 1 | 2430.31963 | 0.00000 |
| frame | 1450.5175 | 2 | 3.27848 | 0.03803 |
| fam | 696.3834 | 1 | 3.14795 | 0.07628 |
| intent_fv_t1gen:frame | 1284.1494 | 2 | 2.90246 | 0.05528 |
| intent_fv_t1gen:fam | 357.6051 | 1 | 1.61653 | 0.20382 |
| frame:fam | 2393.2782 | 2 | 5.40933 | 0.00459 |
| intent_fv_t1gen:frame:fam | 2571.9771 | 2 | 5.81323 | 0.00307 |
| Residuals | 263691.5417 | 1192 | NA | NA |

| | eta.sq | eta.sq.part |
|---|---|---|
| intent_fv_t1gen | 0.54601 | 0.67093 |
| frame | 0.00147 | 0.00547 |
| fam | 0.00071 | 0.00263 |
| intent_fv_t1gen:frame | 0.00130 | 0.00485 |
| intent_fv_t1gen:fam | 0.00036 | 0.00135 |
| frame:fam | 0.00243 | 0.00899 |
| intent_fv_t1gen:frame:fam | 0.00261 | 0.00966 |

#### Contrasts

| | Sum Sq | Df | F value | Pr(>F) |
|---|---|---|---|---|
| (Intercept) | 19226.03563 | 1 | 86.91001 | 0.00000 |
| intent_fv_t1gen | 537629.80664 | 1 | 2430.31963 | 0.00000 |
| fam | 696.38336 | 1 | 3.14795 | 0.07628 |
| cont1 | 1448.02243 | 1 | 6.54569 | 0.01064 |
| cont2 | 0.59441 | 1 | 0.00269 | 0.95867 |
| intent_fv_t1gen:fam | 357.60511 | 1 | 1.61653 | 0.20382 |
| intent_fv_t1gen:cont1 | 1165.73800 | 1 | 5.26964 | 0.02187 |
| fam:cont1 | 2366.58166 | 1 | 10.69797 | 0.00110 |
| intent_fv_t1gen:cont2 | 156.46629 | 1 | 0.70730 | 0.40051 |
| fam:cont2 | 66.55595 | 1 | 0.30086 | 0.58345 |
| intent_fv_t1gen:fam:cont1 | 2566.55624 | 1 | 11.60195 | 0.00068 |
| intent_fv_t1gen:fam:cont2 | 22.64113 | 1 | 0.10235 | 0.74909 |
| Residuals | 263691.54174 | 1192 | NA | NA |

| | eta.sq | eta.sq.part |
|---|---|---|
| intent_fv_t1gen | 0.54601 | 0.67093 |
| fam | 0.00071 | 0.00263 |
| cont1 | 0.00147 | 0.00546 |
| cont2 | 0.00000 | 0.00000 |
| intent_fv_t1gen:fam | 0.00036 | 0.00135 |
| intent_fv_t1gen:cont1 | 0.00118 | 0.00440 |
| fam:cont1 | 0.00240 | 0.00889 |
| intent_fv_t1gen:cont2 | 0.00016 | 0.00059 |
| fam:cont2 | 0.00007 | 0.00025 |
| intent_fv_t1gen:fam:cont1 | 0.00261 | 0.00964 |
| intent_fv_t1gen:fam:cont2 | 0.00002 | 0.00009 |

#### Group Means

| frame | emmean | SE | df | lower.CL | upper.CL |
|---|---|---|---|---|---|
| control | 70.13011 | 0.8042239 | 1192 | 68.55226 | 71.70796 |
| negative | 70.31978 | 0.8238466 | 1192 | 68.70343 | 71.93613 |
| positive | 72.61725 | 0.8013953 | 1192 | 71.04495 | 74.18955 |

| frame | fam | emmean | SE | df | lower.CL | upper.CL |
|---|---|---|---|---|---|---|
| control | unfamiliar | 70.61730 | 1.146919 | 1192 | 68.36709 | 72.86750 |
| negative | unfamiliar | 71.77062 | 1.217889 | 1192 | 69.38117 | 74.16006 |
| positive | unfamiliar | 73.19950 | 1.147633 | 1192 | 70.94790 | 75.45111 |
| control | familiar | 69.64292 | 1.127688 | 1192 | 67.43045 | 71.85540 |
| negative | familiar | 68.86895 | 1.109793 | 1192 | 66.69158 | 71.04631 |
| positive | familiar | 72.03499 | 1.118873 | 1192 | 69.83981 | 74.23017 |

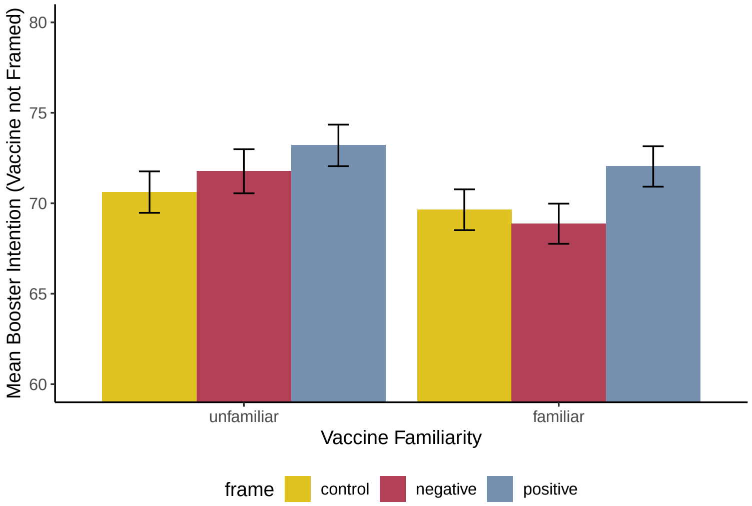


Figure 1.4.1: Graph depicting the Framing * Familiarity interaction for the Booster not framed. Please note that the Unfamiliar condition now refers to those who saw the Moderna vaccine framed, but are rating their intention for the Pfizer vaccine (and vice versa)

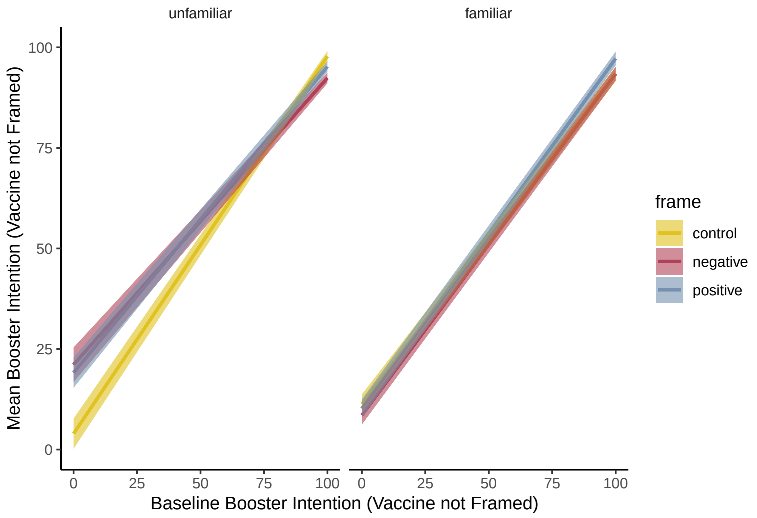


Figure 1.4.2: Graph depicting the Framing * Familiarity * Baseline Intention (for the booster not framed) interaction. Please note that the Unfamiliar condition now refers to those who saw the Moderna vaccine framed, but are rating their intention for the Pfizer vaccine (and vice versa)

### S1.2: STROBE Checklist

STROBE Statement—Checklist of items that should be included in reports of ***cross-sectional studies***

| | Item No | Recommendation | Page No |
|---|---|---|---|
| **Title and abstract** | 1 | (*a*) Indicate the study's design with a commonly used term in the title or the abstract | 1 |
| | | (*b*) Provide in the abstract an informative and balanced summary of what was done and what was found | 1 |
| **Introduction** | | | |
| Background/rationale | 2 | Explain the scientific background and rationale for the investigation being reported | 2 – 3 |
| Objectives | 3 | State specific objectives, including any prespecified hypotheses | 3 |
| **Methods** | | | |
| Study design | 4 | Present key elements of study design early in the paper | 4 - 7 |
| Setting | 5 | Describe the setting, locations, and relevant dates, including periods of recruitment, exposure, follow-up, and data collection | 3 |
| Participants | 6 | (*a*) Give the eligibility criteria, and the sources and methods of selection of participants | 3-4 |
| Variables | 7 | Clearly define all outcomes, exposures, predictors, potential confounders, and effect modifiers. Give diagnostic criteria, if applicable | 4-8 |
| Data sources/ measurement | 8* | For each variable of interest, give sources of data and details of methods of assessment (measurement). Describe comparability of assessment methods if there is more than one group | 4, 6-8 |
| Bias | 9 | Describe any efforts to address potential sources of bias | 4, 9 |
| Study size | 10 | Explain how the study size was arrived at | 9 |
| Quantitative variables | 11 | Explain how quantitative variables were handled in the analyses. If applicable, describe which groupings were chosen and why | 9 |
| Statistical methods | 12 | (*a*) Describe all statistical methods, including those used to control for confounding | 9 |
| | | (*b*) Describe any methods used to examine subgroups and interactions | 9 |
| | | (*c*) Explain how missing data were addressed | N/A (see page 4) |
| | | (*d*) If applicable, describe analytical methods taking account of sampling strategy | N/A |
| | | (*e*) Describe any sensitivity analyses | N/A |
| **Results** | | | |
| Participants | 13* | (a) Report numbers of individuals at each stage of study—eg numbers potentially eligible, examined for eligibility, confirmed eligible, included in the study, completing follow-up, and analysed | 9 |
| | | (b) Give reasons for non-participation at each stage | 9 |
| | | (c) Consider use of a flow diagram | - |
| Descriptive data | 14* | (a) Give characteristics of study participants (eg demographic, clinical, social) and information on exposures and potential confounders | 9-11 |
| | | (b) Indicate number of participants with missing data for each variable of interest | N/A (see page 4) |

| | | | |
|---|---|---|---|
| Outcome data | 15* | Report numbers of outcome events or summary measures | 13 |
| Main results | 16 | (*a*) Give unadjusted estimates and, if applicable, confounder-adjusted estimates and their precision (eg, 95% confidence interval). Make clear which confounders were adjusted for and why they were included | 12-13 |
| | | (*b*) Report category boundaries when continuous variables were categorized | N/A |
| | | (*c*) If relevant, consider translating estimates of relative risk into absolute risk for a meaningful time period | N/A |
| Other analyses | 17 | Report other analyses done—eg analyses of subgroups and interactions, and sensitivity analyses | N/A |
| **Discussion** | | | |
| Key results | 18 | Summarise key results with reference to study objectives | 14-15 |
| Limitations | 19 | Discuss limitations of the study, taking into account sources of potential bias or imprecision. Discuss both direction and magnitude of any potential bias | 16-17 |
| Interpretation | 20 | Give a cautious overall interpretation of results considering objectives, limitations, multiplicity of analyses, results from similar studies, and other relevant evidence | 17 |
| Generalisability | 21 | Discuss the generalisability (external validity) of the study results | 17 |
| **Other information** | | | |
| Funding | 22 | Give the source of funding and the role of the funders for the present study and, if applicable, for the original study on which the present article is based | 18 |

*Give information separately for exposed and unexposed groups.

**Note:** An Explanation and Elaboration article discusses each checklist item and gives methodological background and published examples of transparent reporting. The STROBE checklist is best used in conjunction with this article (freely available on the Web sites of PLoS Medicine at http://www.plosmedicine.org/, Annals of Internal Medicine at http://www.annals.org/, and Epidemiology at http://www.epidem.com/). Information on the STROBE Initiative is available at www.strobe-statement.org.

### S1.3: Survey Structure

The items included in the current study are presented on the following pages. The Familiar (i.e., Pfizer) / Negative Framing Condition is used as the example. Responses are dummy data for illustrative purposes.

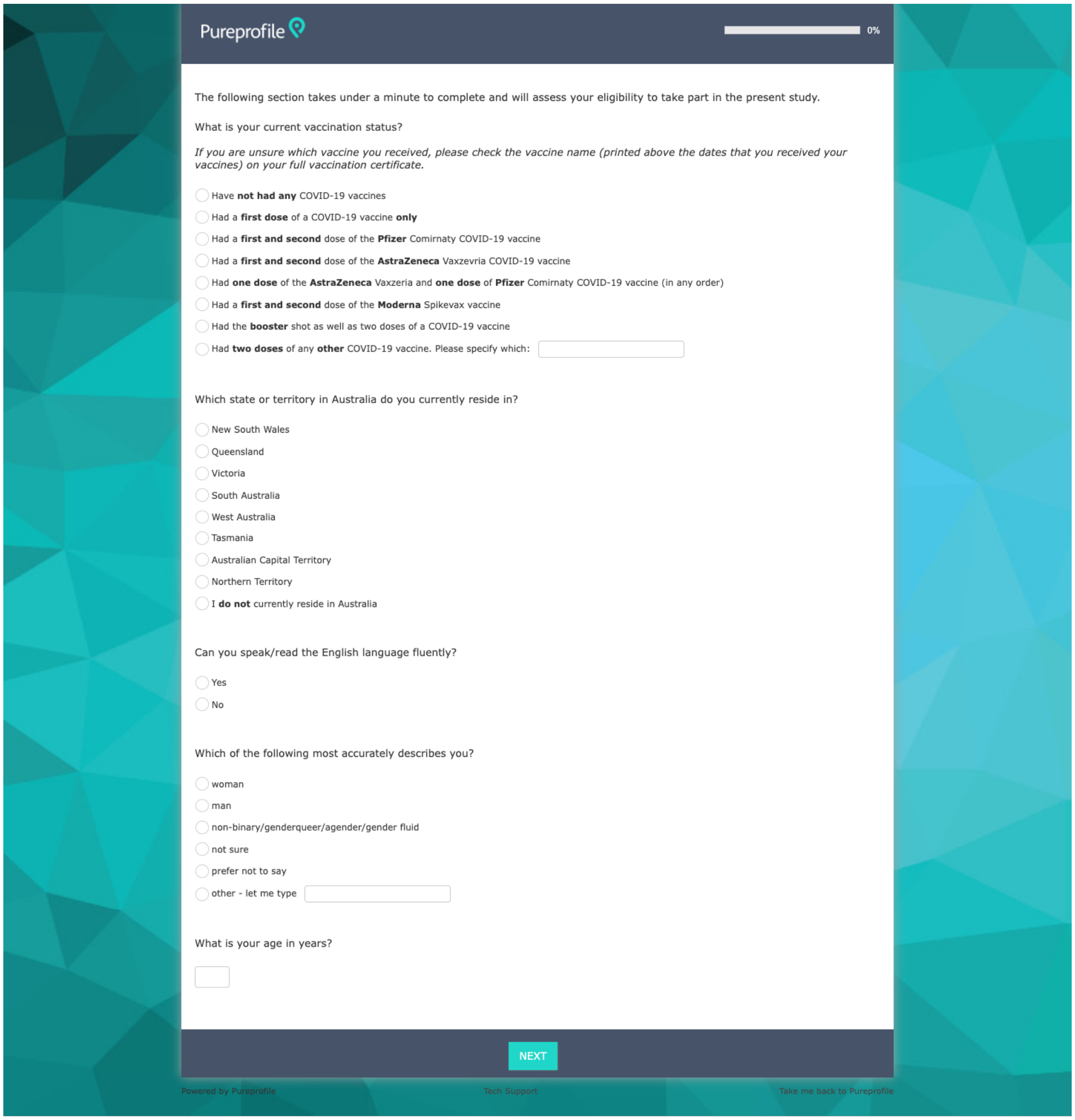
Pureprofile

0%

The following section takes under a minute to complete and will assess your eligibility to take part in the present study.

What is your current vaccination status?

*If you are unsure which vaccine you received, please check the vaccine name (printed above the dates that you received your vaccines) on your full vaccination certificate.*

- Have **not had any** COVID-19 vaccines
- Had a **first dose** of a COVID-19 vaccine **only**
- Had a **first and second** dose of the **Pfizer** Comirnaty COVID-19 vaccine
- Had a **first and second** dose of the **AstraZeneca** Vaxzevria COVID-19 vaccine
- Had **one dose** of the **AstraZeneca** Vaxzeria and **one dose** of **Pfizer** Comirnaty COVID-19 vaccine (in any order)
- Had a **first and second** dose of the **Moderna** Spikevax vaccine
- Had the **booster** shot as well as two doses of a COVID-19 vaccine
- Had **two doses** of any **other** COVID-19 vaccine. Please specify which:

Which state or territory in Australia do you currently reside in?

- New South Wales
- Queensland
- Victoria
- South Australia
- West Australia
- Tasmania
- Australian Capital Territory
- Northern Territory
- I **do not** currently reside in Australia

Can you speak/read the English language fluently?

- Yes
- No

Which of the following most accurately describes you?

- woman
- man
- non-binary/genderqueer/agender/gender fluid
- not sure
- prefer not to say
- other - let me type

What is your age in years?

NEXT

Powered by Pureprofile Tech Support Take me back to Pureprofile

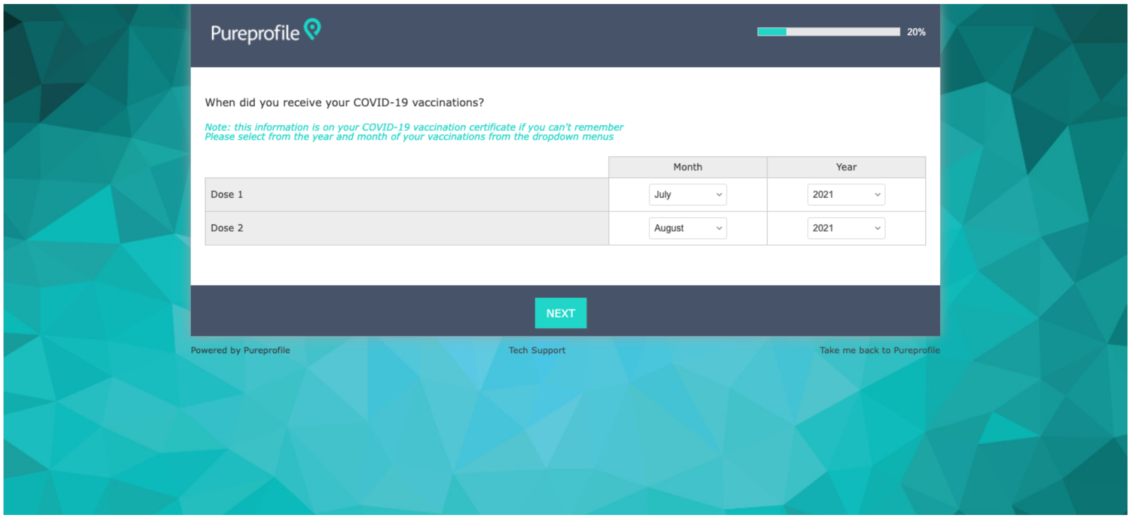
Pureprofile
20%
When did you receive your COVID-19 vaccinations?
Note: this information is on your COVID-19 vaccination certificate if you can't remember
Please select from the year and month of your vaccinations from the dropdown menus
Month
Year
Dose 1
July
2021
Dose 2
August
2021
NEXT
Powered by Pureprofile
Tech Support
Take me back to Pureprofile

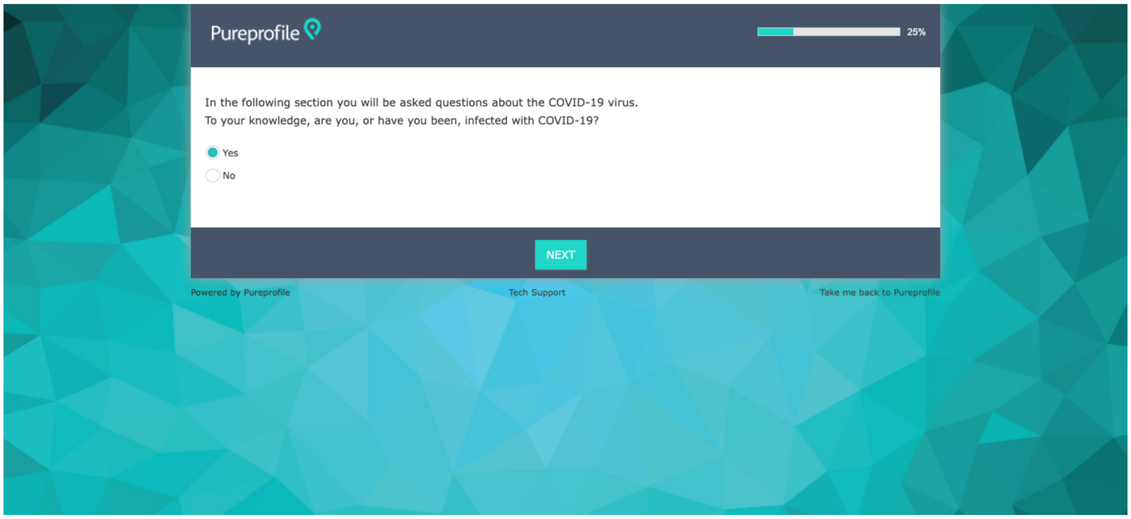
Pureprofile
25%
In the following section you will be asked questions about the COVID-19 virus.
To your knowledge, are you, or have you been, infected with COVID-19?
Yes
No
NEXT
Powered by Pureprofile
Tech Support
Take me back to Pureprofile

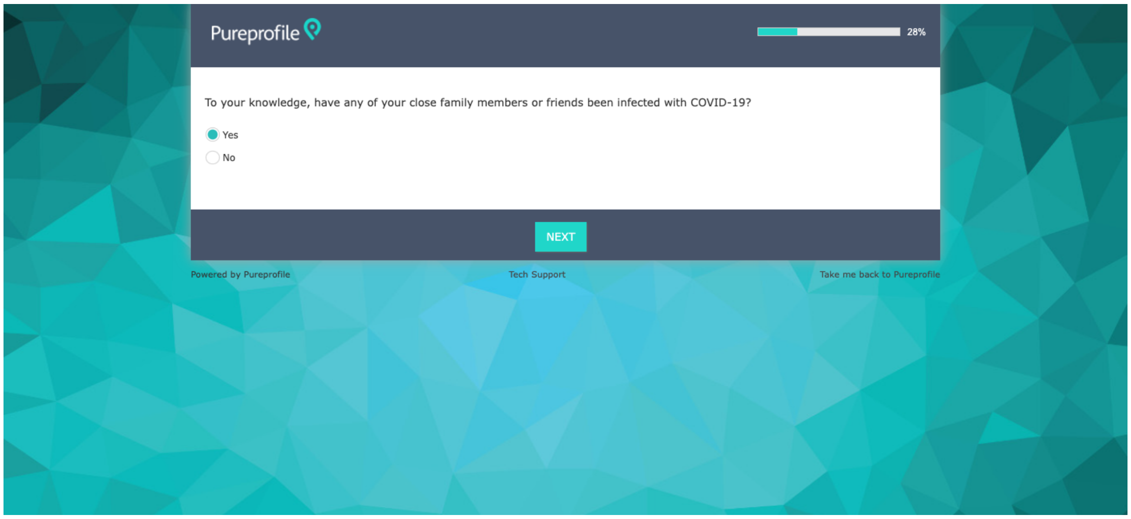
Pureprofile
28%
To your knowledge, have any of your close family members or friends been infected with COVID-19?
Yes
No
NEXT
Powered by Pureprofile
Tech Support
Take me back to Pureprofile

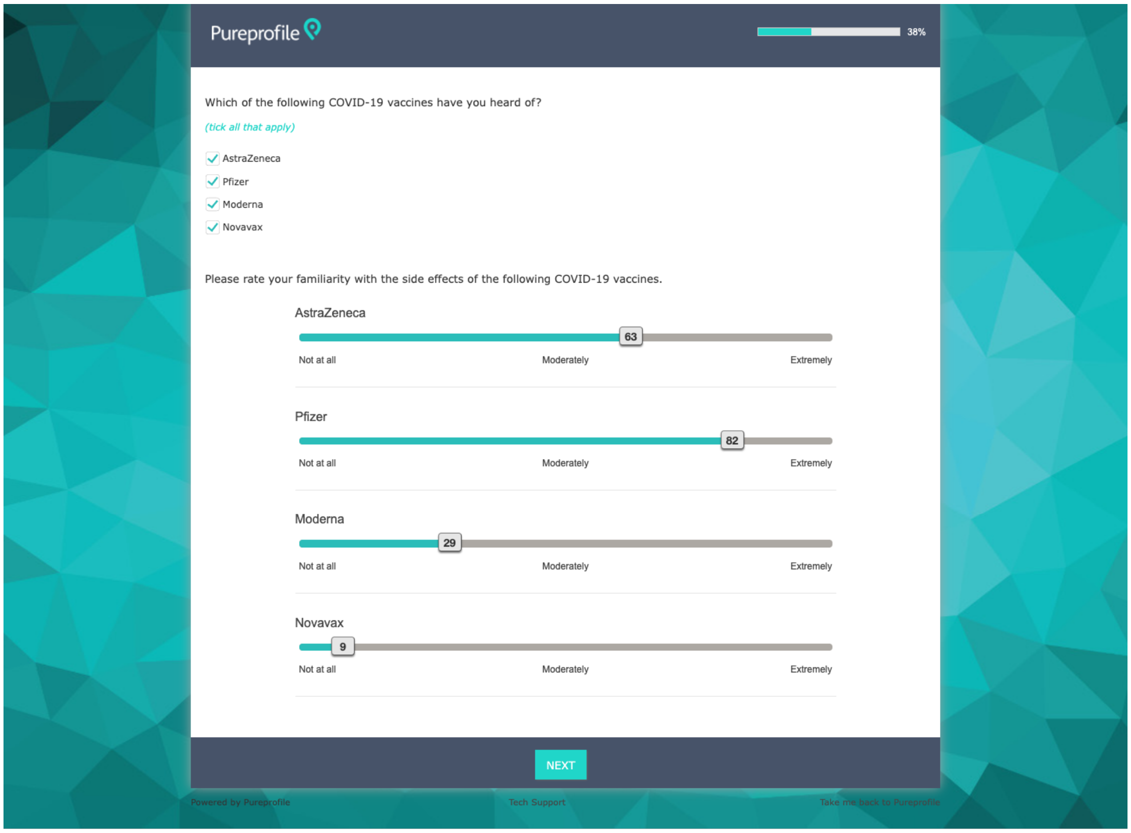
Pureprofile
38%
Which of the following COVID-19 vaccines have you heard of?
(tick all that apply)
AstraZeneca
Pfizer
Moderna
Novavax
Please rate your familiarity with the side effects of the following COVID-19 vaccines.
AstraZeneca
63
Not at all
Moderately
Extremely
Pfizer
82
Not at all
Moderately
Extremely
Moderna
29
Not at all
Moderately
Extremely
Novavax
9
Not at all
Moderately
Extremely
NEXT
Powered by Pureprofile
Tech Support
Take me back to Pureprofile

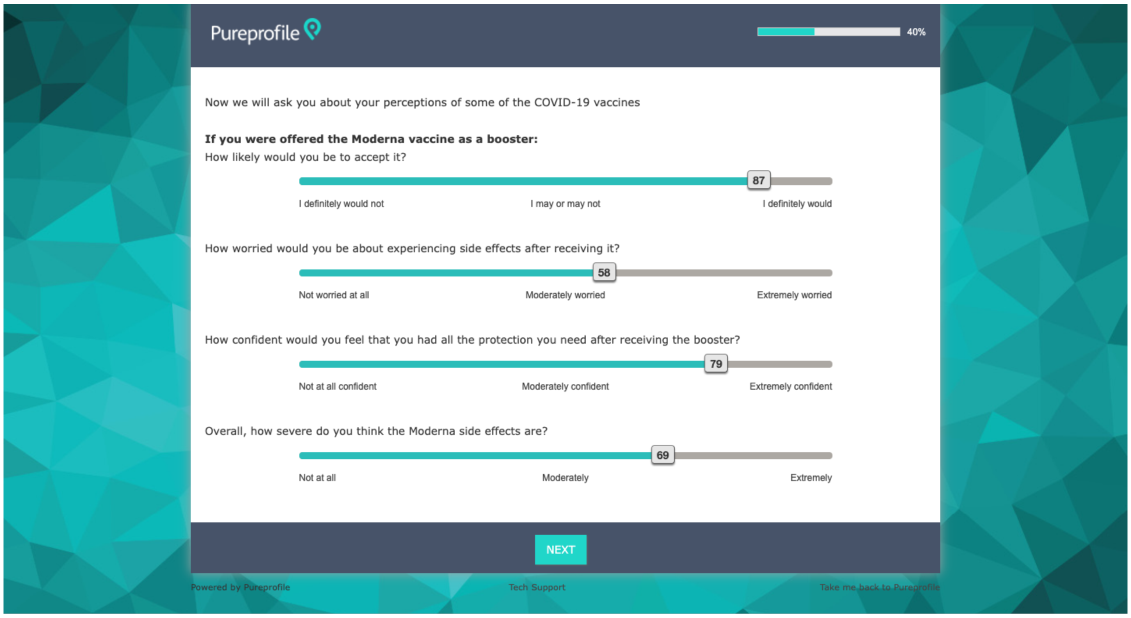
Pureprofile
40%
Now we will ask you about your perceptions of some of the COVID-19 vaccines
If you were offered the Moderna vaccine as a booster:
How likely would you be to accept it?
87
I definitely would not
I may or may not
I definitely would
How worried would you be about experiencing side effects after receiving it?
58
Not worried at all
Moderately worried
Extremely worried
How confident would you feel that you had all the protection you need after receiving the booster?
79
Not at all confident
Moderately confident
Extremely confident
Overall, how severe do you think the Moderna side effects are?
69
Not at all
Moderately
Extremely
NEXT
Powered by Pureprofile
Tech Support
Take me back to Pureprofile

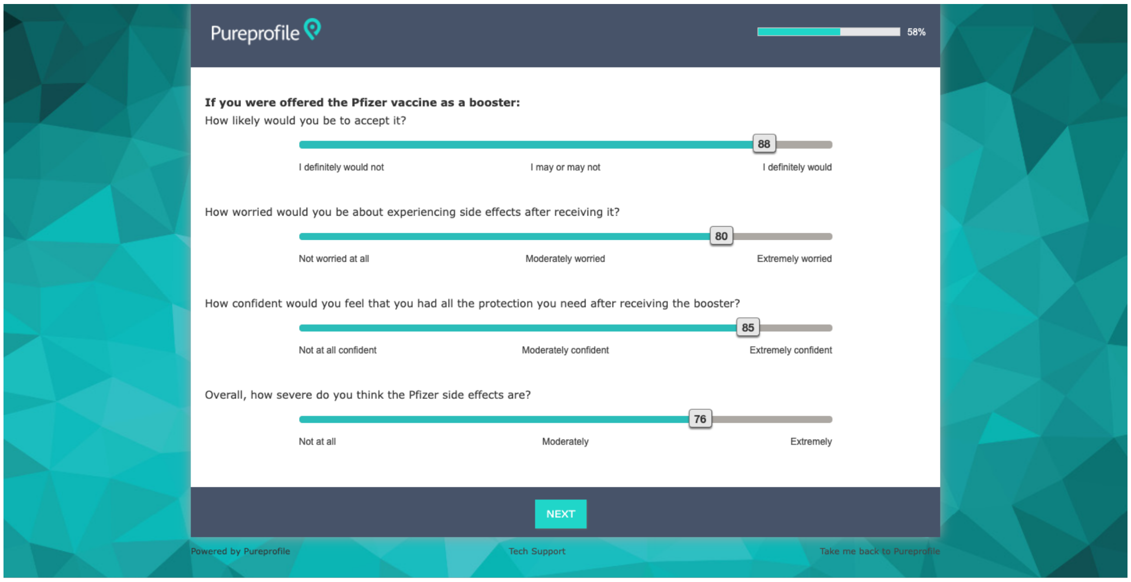
Pureprofile
58%
If you were offered the Pfizer vaccine as a booster:
How likely would you be to accept it?
88
I definitely would not
I may or may not
I definitely would
How worried would you be about experiencing side effects after receiving it?
80
Not worried at all
Moderately worried
Extremely worried
How confident would you feel that you had all the protection you need after receiving the booster?
85
Not at all confident
Moderately confident
Extremely confident
Overall, how severe do you think the Pfizer side effects are?
76
Not at all
Moderately
Extremely
NEXT
Powered by Pureprofile
Tech Support
Take me back to Pureprofile

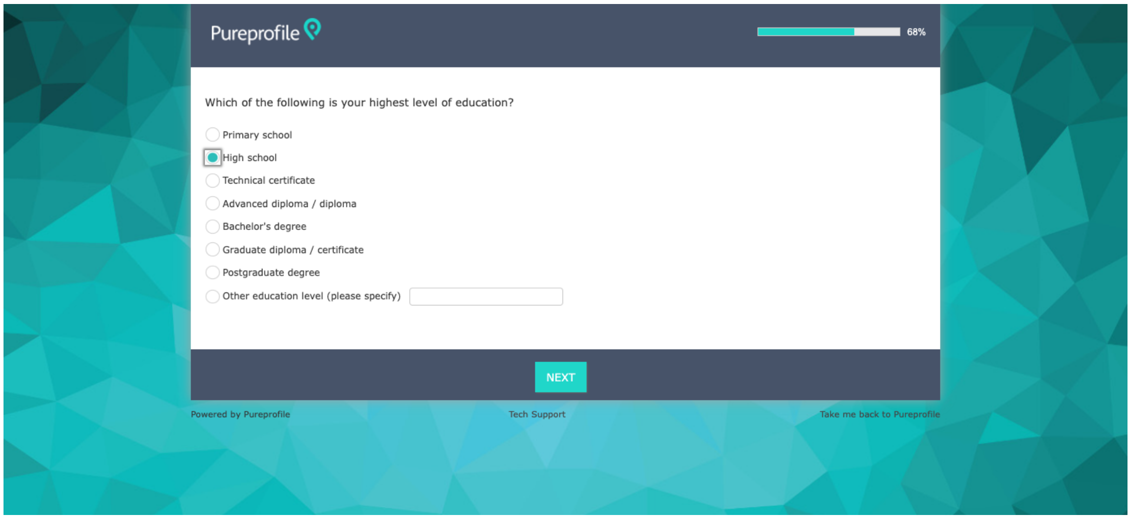
Pureprofile
68%
Which of the following is your highest level of education?
Primary school
High school
Technical certificate
Advanced diploma / diploma
Bachelor's degree
Graduate diploma / certificate
Postgraduate degree
Other education level (please specify)
NEXT
Powered by Pureprofile
Tech Support
Take me back to Pureprofile

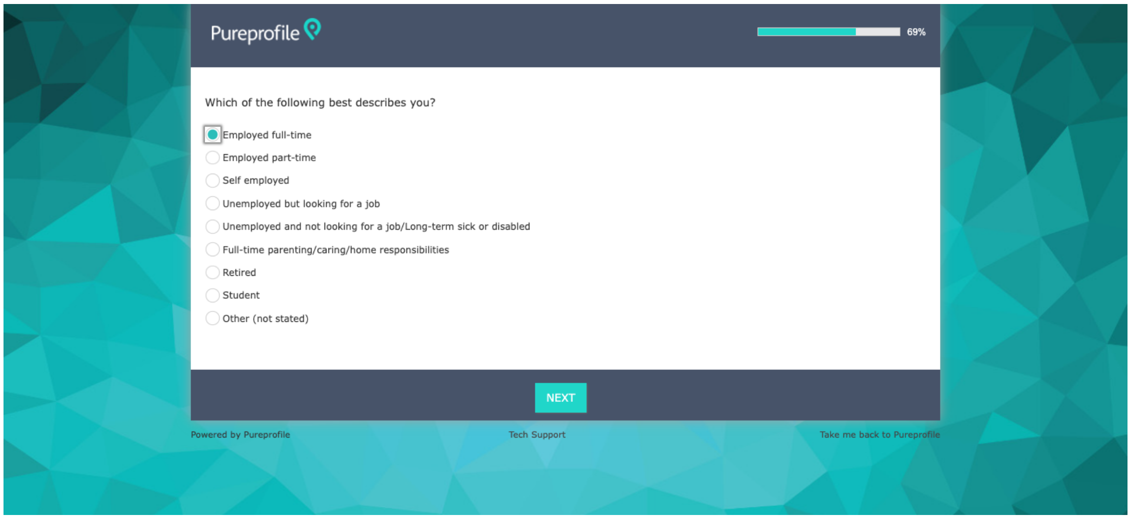
Pureprofile
69%
Which of the following best describes you?
Employed full-time
Employed part-time
Self employed
Unemployed but looking for a job
Unemployed and not looking for a job/Long-term sick or disabled
Full-time parenting/caring/home responsibilities
Retired
Student
Other (not stated)
NEXT
Powered by Pureprofile
Tech Support
Take me back to Pureprofile

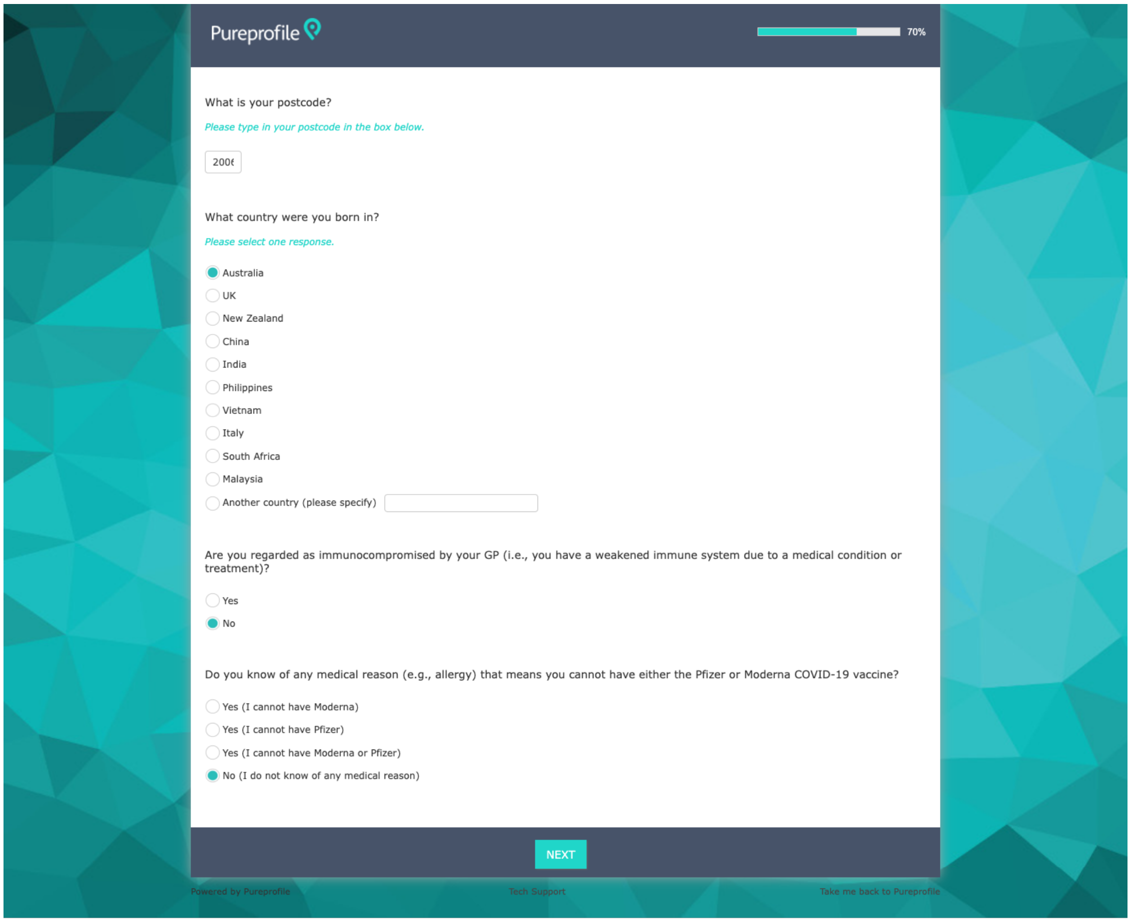

Pureprofile 70%

What is your postcode?

*Please type in your postcode in the box below.*

2006

What country were you born in?

*Please select one response.*

- (●) Australia
- ( ) UK
- ( ) New Zealand
- ( ) China
- ( ) India
- ( ) Philippines
- ( ) Vietnam
- ( ) Italy
- ( ) South Africa
- ( ) Malaysia
- ( ) Another country (please specify)

Are you regarded as immunocompromised by your GP (i.e., you have a weakened immune system due to a medical condition or treatment)?

- ( ) Yes
- (●) No

Do you know of any medical reason (e.g., allergy) that means you cannot have either the Pfizer or Moderna COVID-19 vaccine?

- ( ) Yes (I cannot have Moderna)
- ( ) Yes (I cannot have Pfizer)
- ( ) Yes (I cannot have Moderna or Pfizer)
- (●) No (I do not know of any medical reason)

NEXT

Powered by Pureprofile Tech Support Take me back to Pureprofile

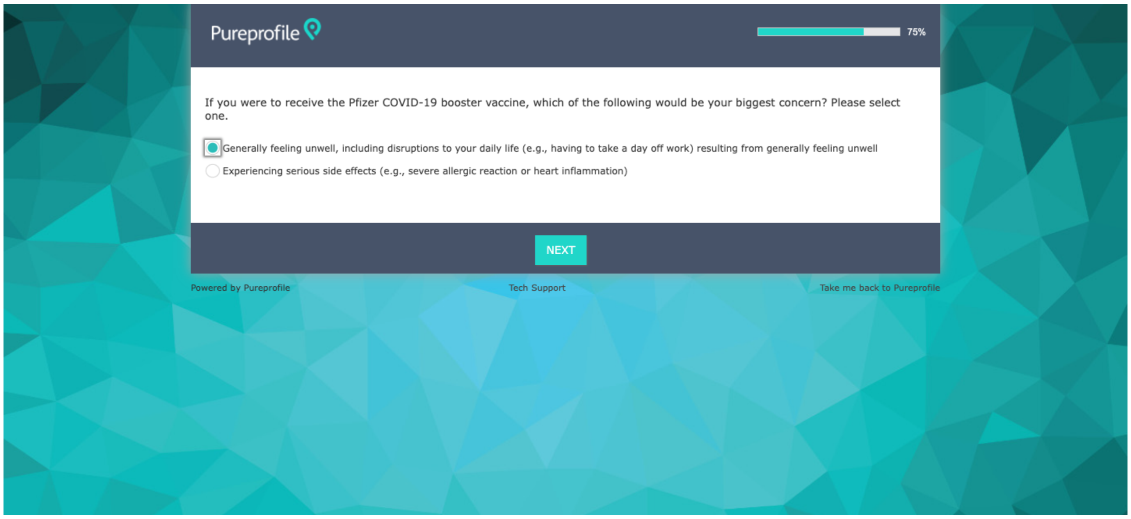

Pureprofile 75%

If you were to receive the Pfizer COVID-19 booster vaccine, which of the following would be your biggest concern? Please select one.

- (●) Generally feeling unwell, including disruptions to your daily life (e.g., having to take a day off work) resulting from generally feeling unwell
- ( ) Experiencing serious side effects (e.g., severe allergic reaction or heart inflammation)

NEXT

Powered by Pureprofile Tech Support Take me back to Pureprofile

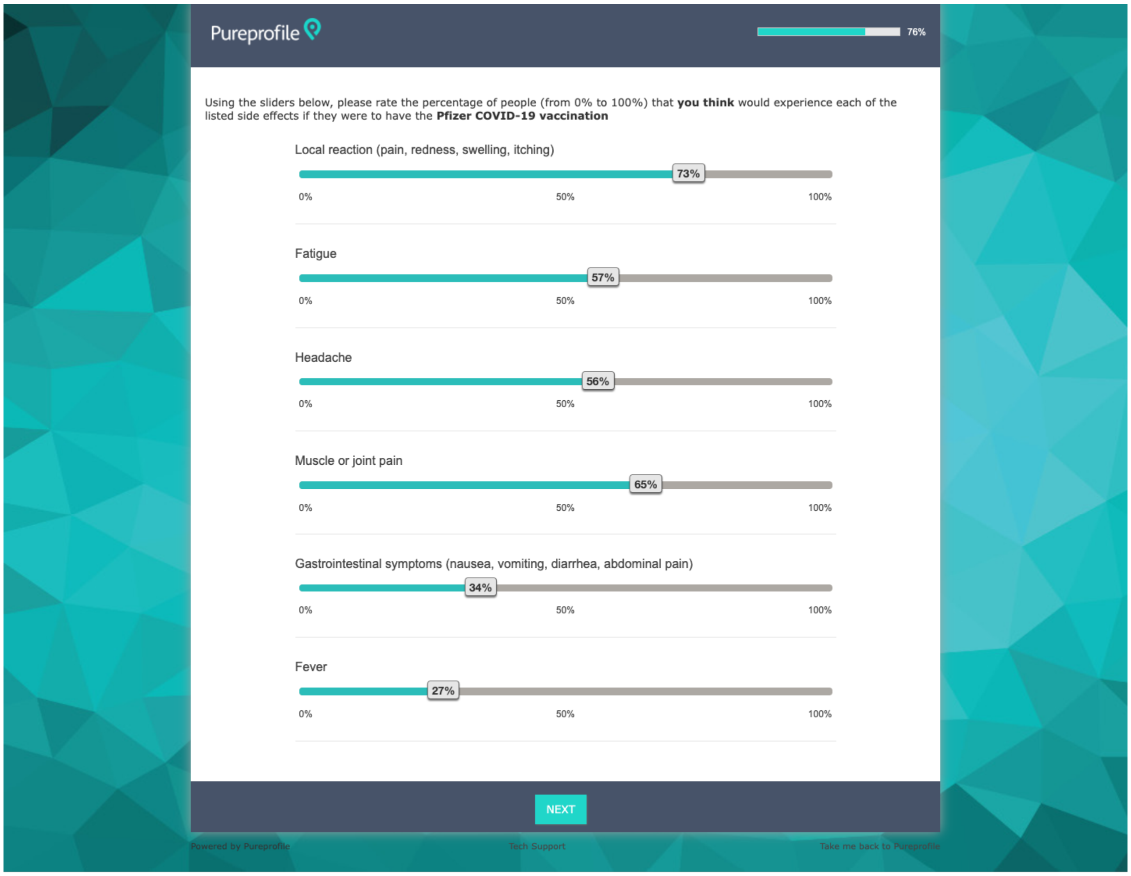
Pureprofile
76%
Using the sliders below, please rate the percentage of people (from 0% to 100%) that you think would experience each of the listed side effects if they were to have the Pfizer COVID-19 vaccination
Local reaction (pain, redness, swelling, itching)
73%
0%
50%
100%
Fatigue
57%
Headache
56%
Muscle or joint pain
65%
Gastrointestinal symptoms (nausea, vomiting, diarrhea, abdominal pain)
34%
Fever
27%
NEXT
Powered by Pureprofile
Tech Support
Take me back to Pureprofile

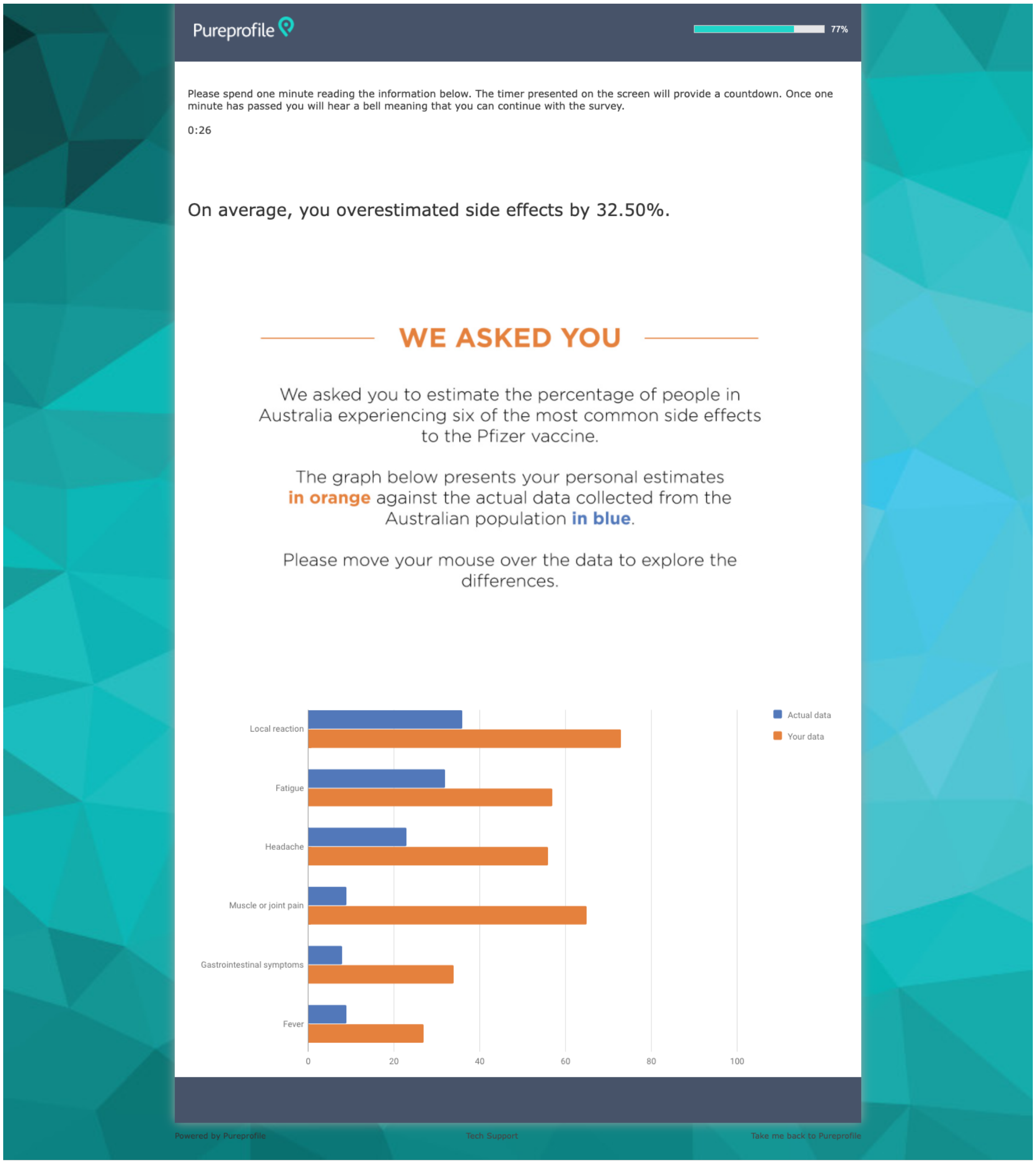
Pureprofile
77%
Please spend one minute reading the information below. The timer presented on the screen will provide a countdown. Once one minute has passed you will hear a bell meaning that you can continue with the survey.
0:26
On average, you overestimated side effects by 32.50%.
WE ASKED YOU
We asked you to estimate the percentage of people in Australia experiencing six of the most common side effects to the Pfizer vaccine.
The graph below presents your personal estimates in orange against the actual data collected from the Australian population in blue.
Please move your mouse over the data to explore the differences.
Local reaction
Fatigue
Headache
Muscle or joint pain
Gastrointestinal symptoms
Fever
0
20
40
60
80
100
Actual data
Your data
Powered by Pureprofile
Tech Support
Take me back to Pureprofile

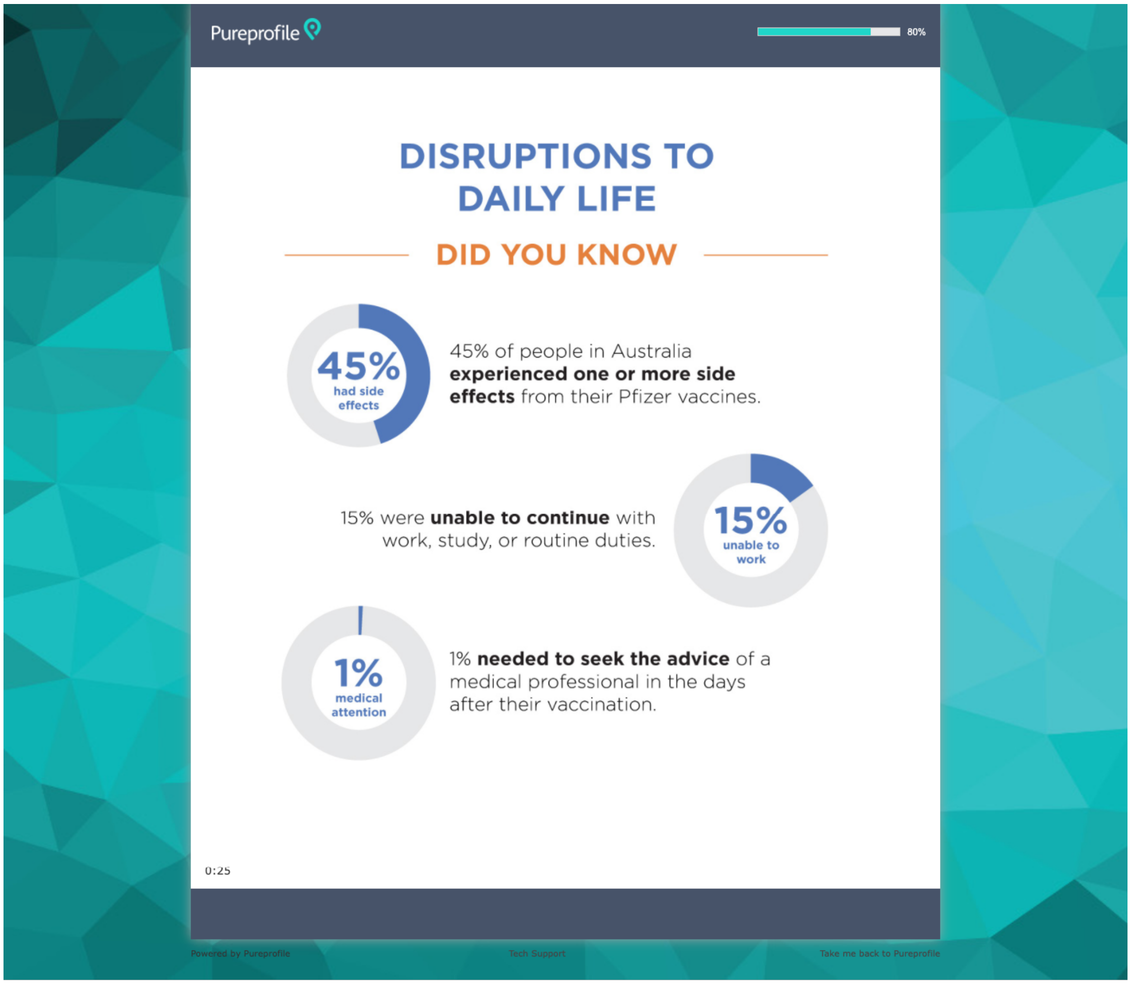
Pureprofile
80%
DISRUPTIONS TO DAILY LIFE
DID YOU KNOW
45% had side effects
45% of people in Australia experienced one or more side effects from their Pfizer vaccines.
15% were unable to continue with work, study, or routine duties.
15% unable to work
1% medical attention
1% needed to seek the advice of a medical professional in the days after their vaccination.
0:25
Powered by Pureprofile
Tech Support
Take me back to Pureprofile

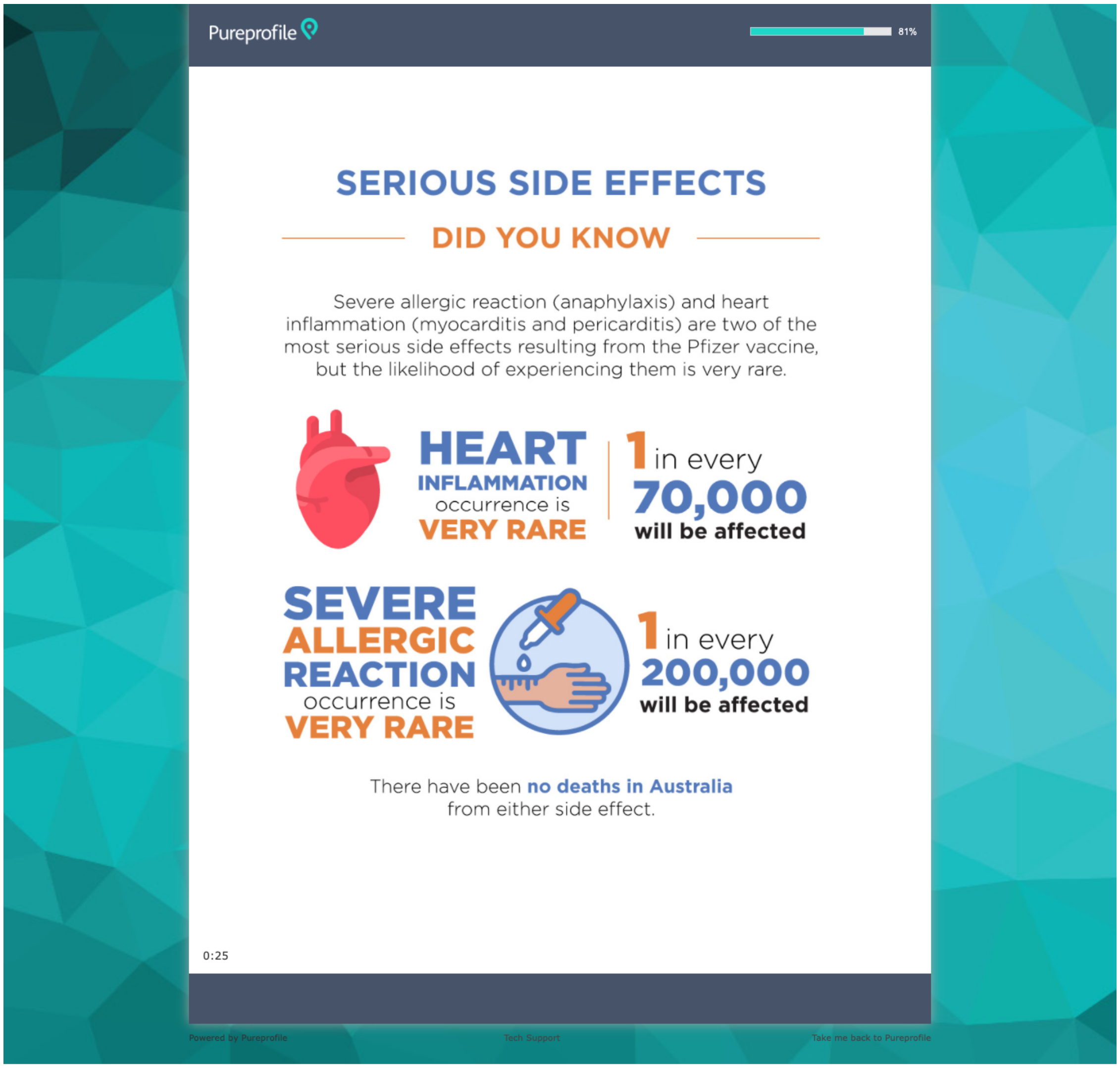
Pureprofile
81%
SERIOUS SIDE EFFECTS
DID YOU KNOW
Severe allergic reaction (anaphylaxis) and heart inflammation (myocarditis and pericarditis) are two of the most serious side effects resulting from the Pfizer vaccine, but the likelihood of experiencing them is very rare.
HEART INFLAMMATION occurrence is VERY RARE
1 in every 70,000 will be affected
SEVERE ALLERGIC REACTION occurrence is VERY RARE
1 in every 200,000 will be affected
There have been no deaths in Australia from either side effect.
0:25

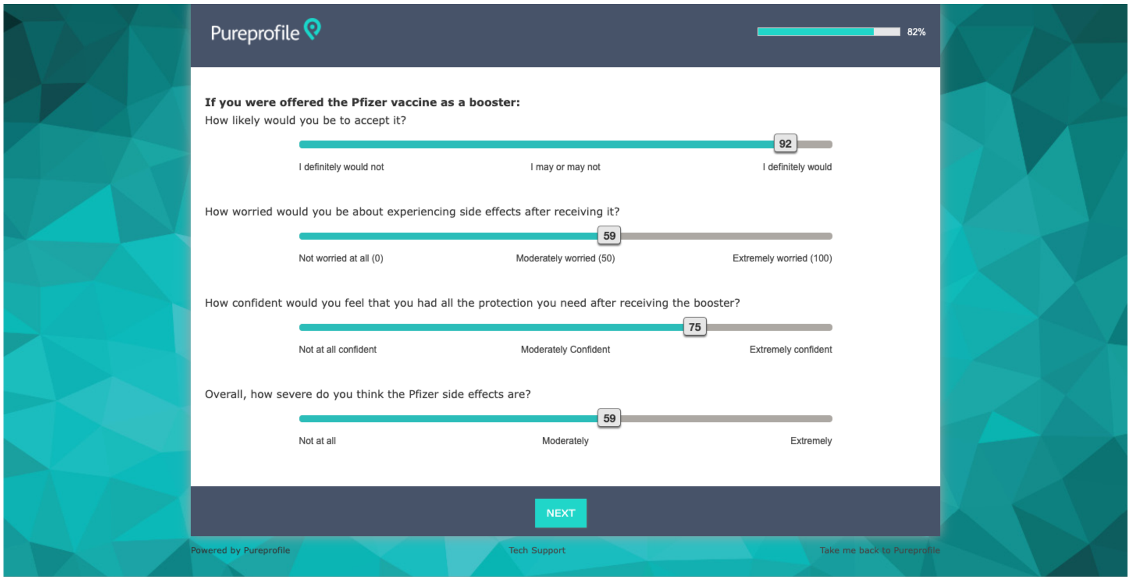
Pureprofile
82%
If you were offered the Pfizer vaccine as a booster:
How likely would you be to accept it?
92
I definitely would not
I may or may not
I definitely would
How worried would you be about experiencing side effects after receiving it?
59
Not worried at all (0)
Moderately worried (50)
Extremely worried (100)
How confident would you feel that you had all the protection you need after receiving the booster?
75
Not at all confident
Moderately Confident
Extremely confident
Overall, how severe do you think the Pfizer side effects are?
59
Not at all
Moderately
Extremely
NEXT
Powered by Pureprofile
Tech Support
Take me back to Pureprofile

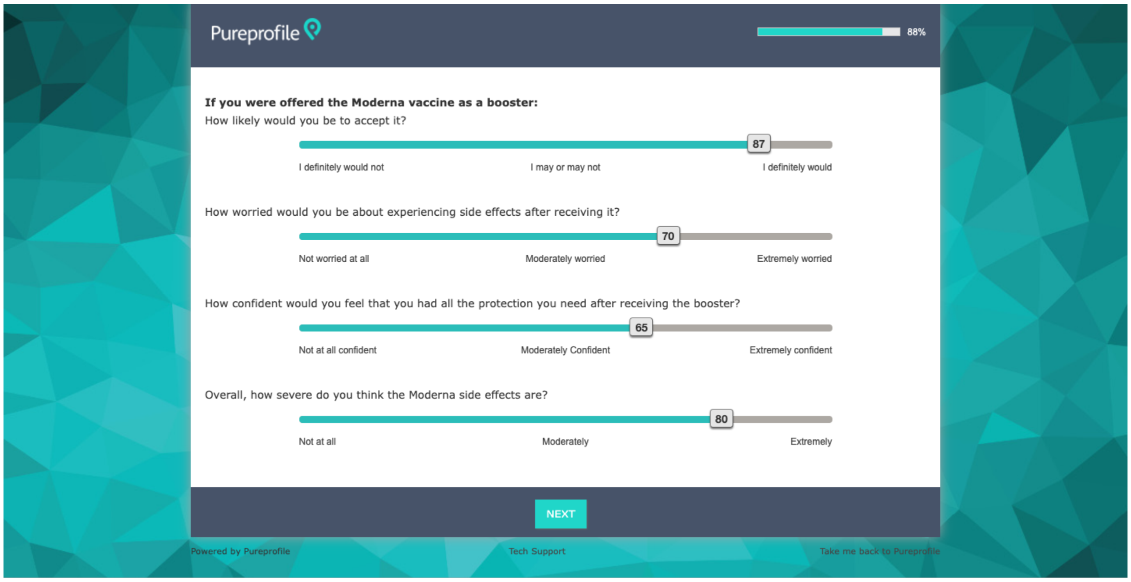
Pureprofile
88%
If you were offered the Moderna vaccine as a booster:
How likely would you be to accept it?
87
I definitely would not
I may or may not
I definitely would
How worried would you be about experiencing side effects after receiving it?
70
Not worried at all
Moderately worried
Extremely worried
How confident would you feel that you had all the protection you need after receiving the booster?
65
Not at all confident
Moderately Confident
Extremely confident
Overall, how severe do you think the Moderna side effects are?
80
Not at all
Moderately
Extremely
NEXT
Powered by Pureprofile
Tech Support
Take me back to Pureprofile

#### S1.4: Infographics used in the study

##### Negative Framing - Infographics.

###### Pfizer: Side Effect Distruptions

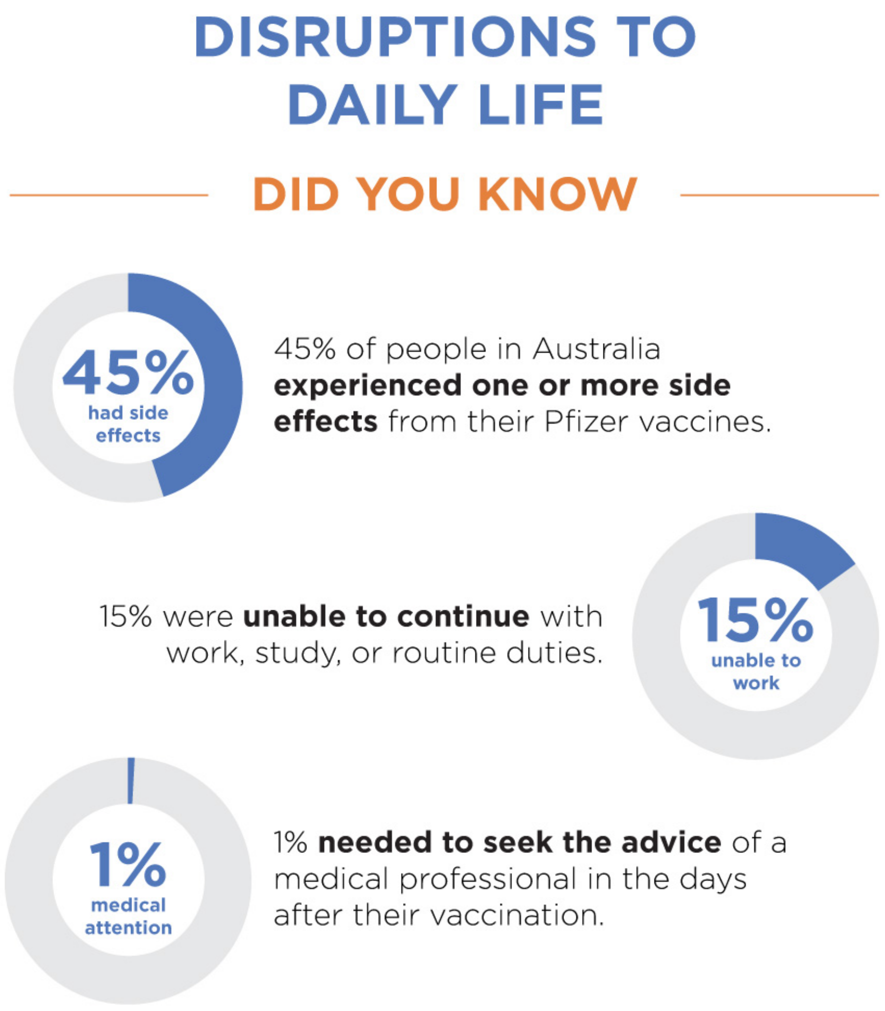

**Pfizer: Sever Side Effects**

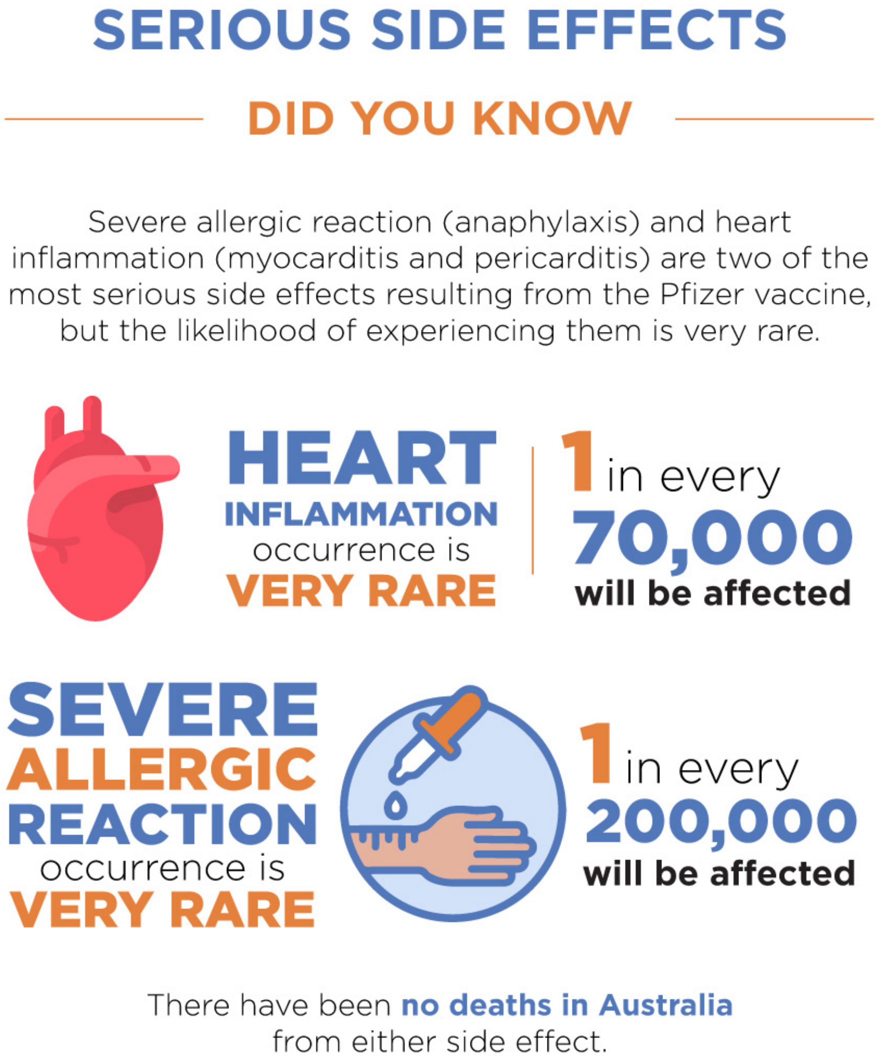

**Pfizer: Side Effect Distruptions**

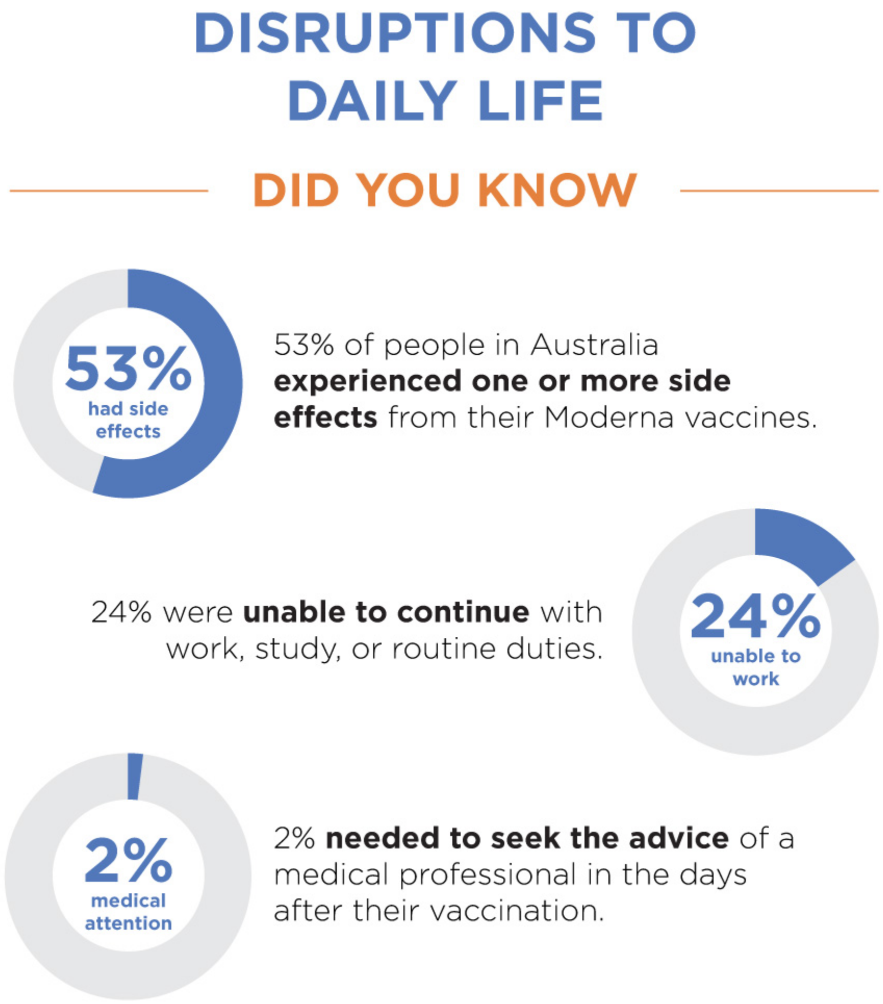

**Pfizer: Severe Side Effects**

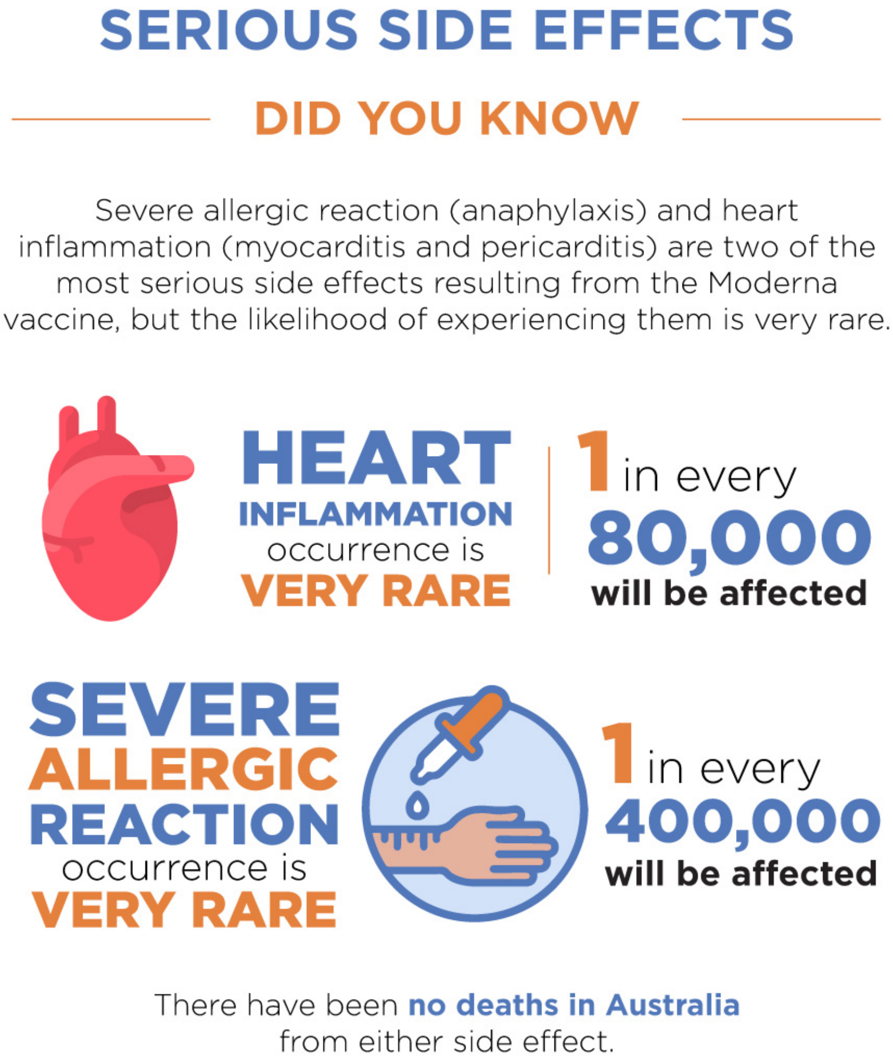

#### Positive Framing - Infographics.

##### Pfizer: Side Effect Distruptions

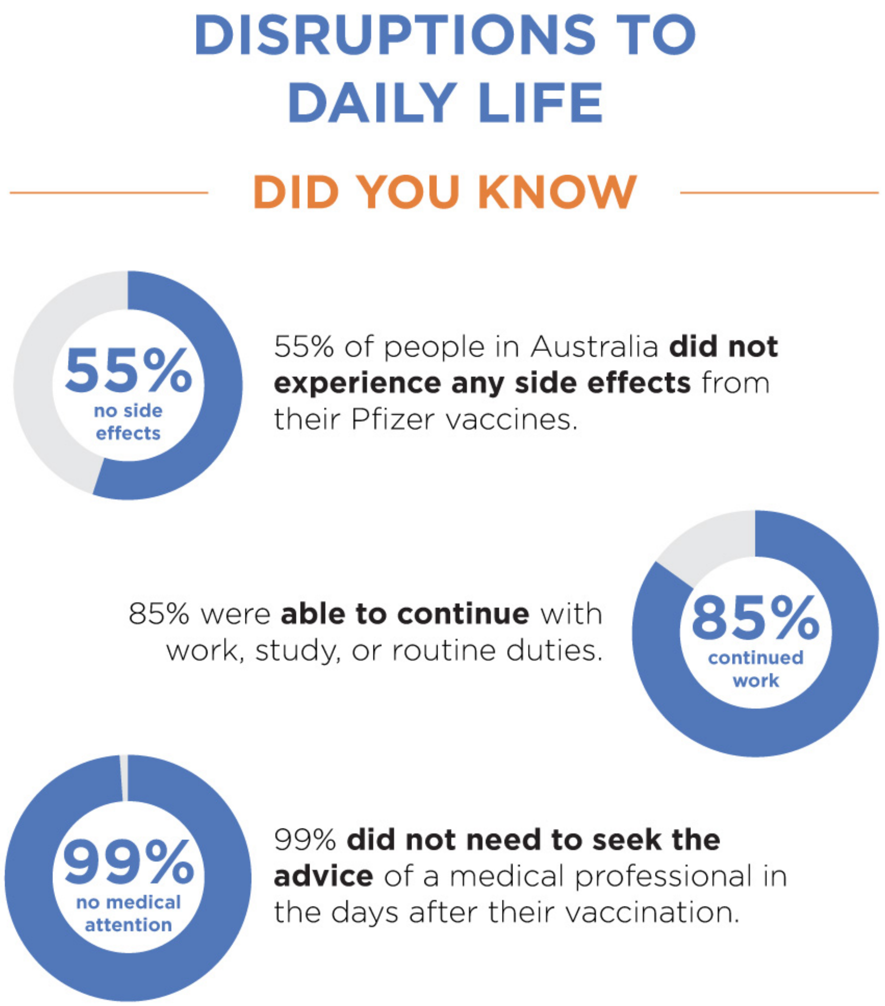

**Pfizer: Severe Side Effects**

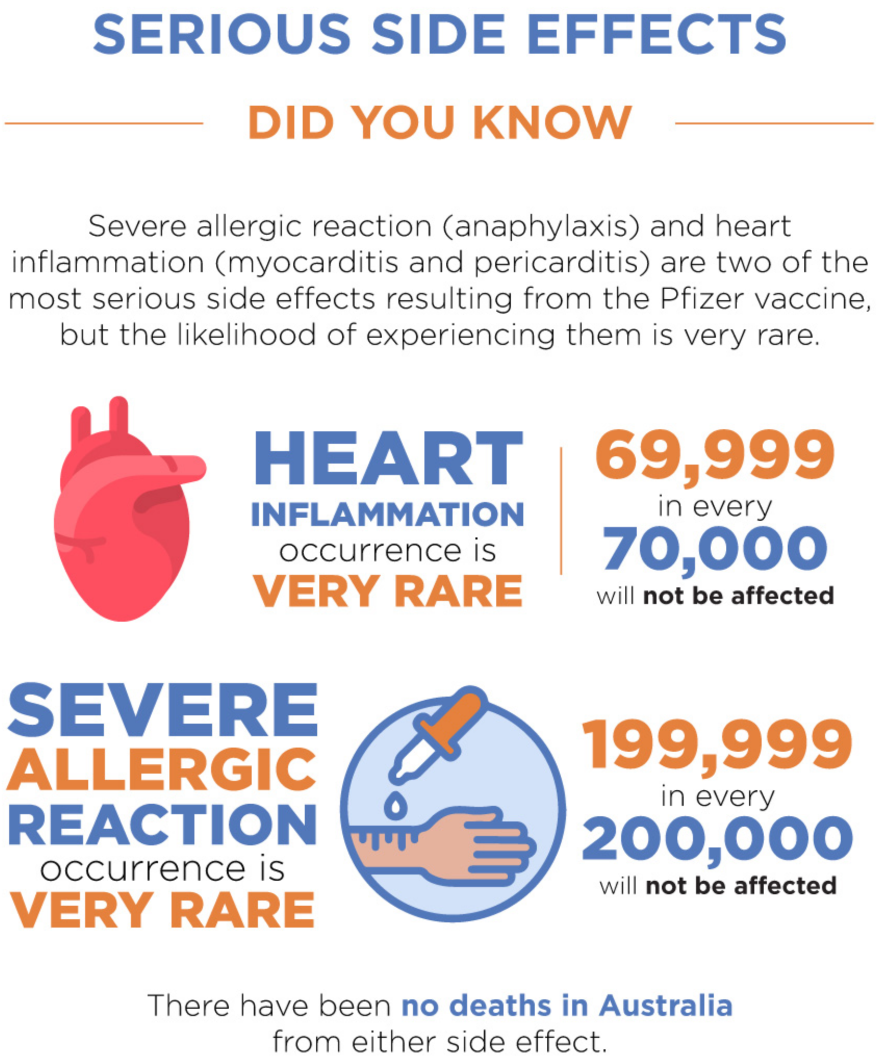

**Pfizer: Side Effect Distruptions**

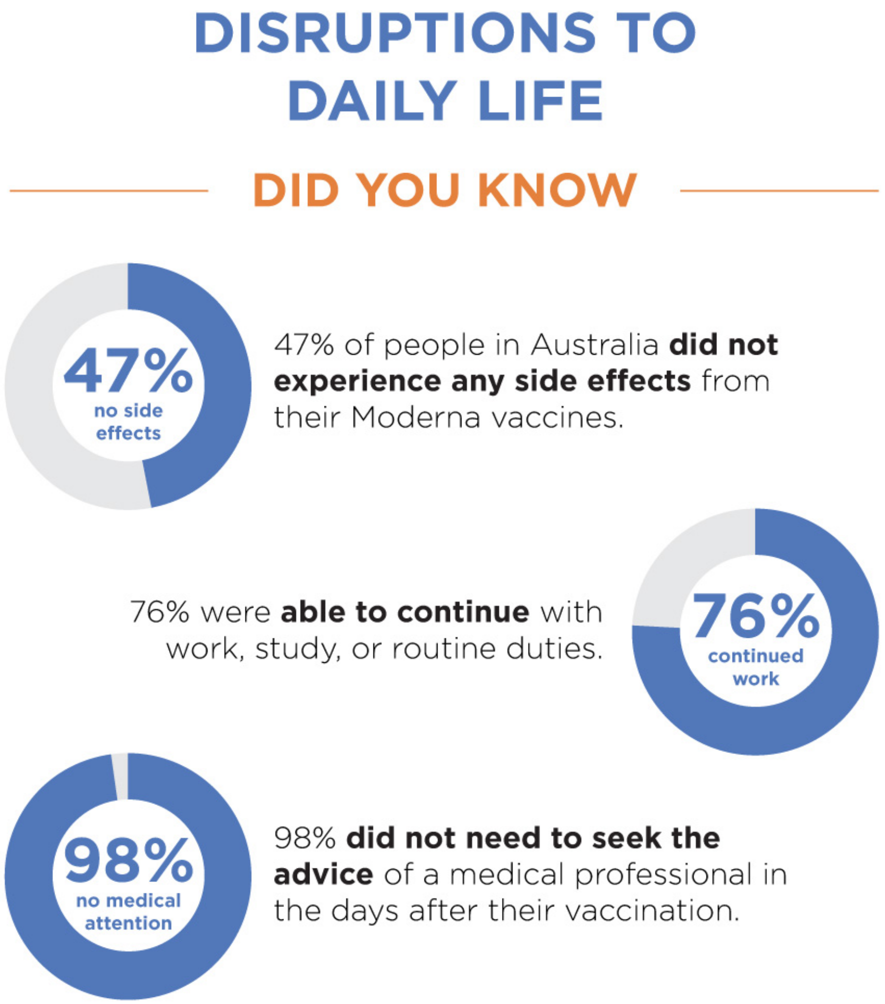

**Pfizer: Severe Side Effects**

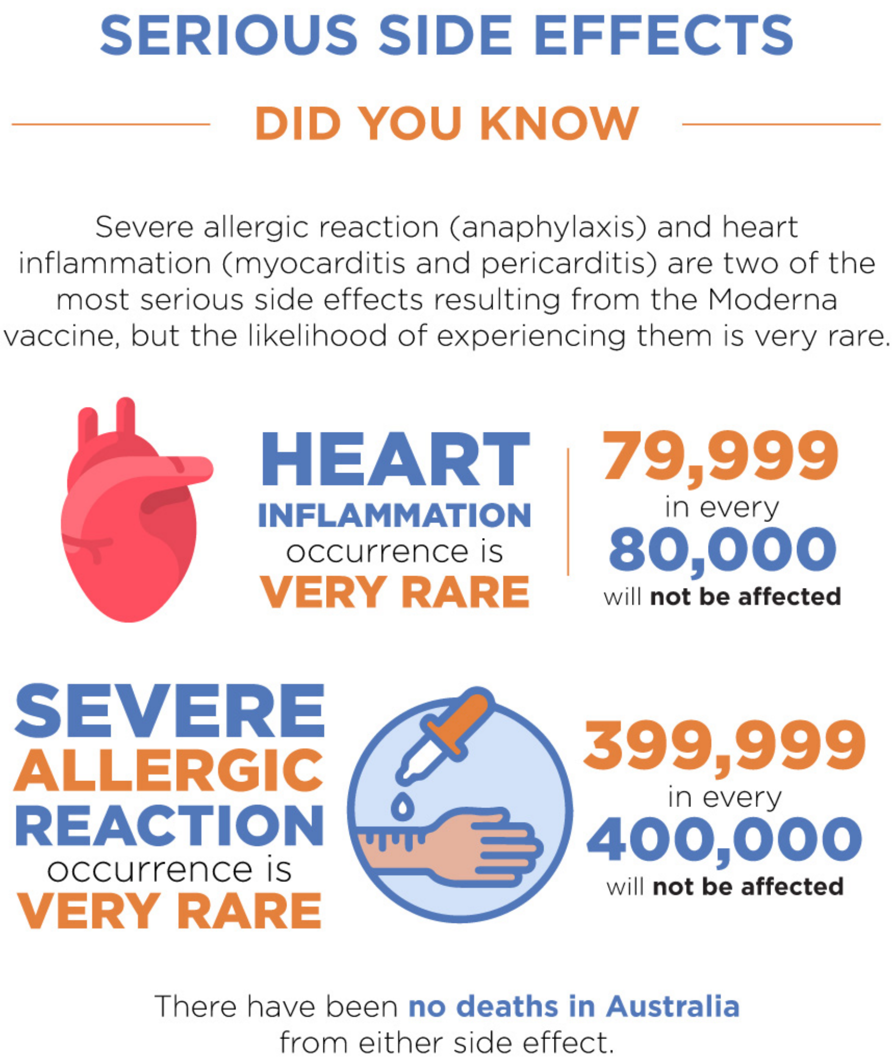

#### S1.5: Pre-analysis data cleaning

**Participants (N = 138) were excluded based on meeting one or more of the following pre-registered quality control checks:**

1. Completed the study quicker than would be expected given a reasonable reading rate (average length of survey/3)
2. Failed to answer attention questions appropriately (e.g., place slider in X position)
3. Stating that they had not received a booster vaccine at screening, then identifying that they had during the survey (i.e., inconsistent answers)

#### S1.6: Primary Analysis - Full Statistical Model

**Item wording:**

"If you were offered the [framed vaccine] as a booster how likely would you be to accept it?

**Variable names:**

- intent_t2 (intention to receive the framed booster vaccine post-intervention)
- intent_fv_t1 (intention to receive the framed booster vaccine at baseline)

##### Overall effects

| | Sum Sq | Df | F value | Pr(>F) |
|---|---|---|---|---|
| (Intercept) | 40226.2838 | 1 | 147.47048 | 0.00000 |
| intent_fv_t1 | 468588.8340 | 1 | 1717.85743 | 0.00000 |
| frame | 4546.6762 | 2 | 8.33411 | 0.00025 |
| fam | 243.7821 | 1 | 0.89371 | 0.34467 |
| intent_fv_t1:frame | 2708.3866 | 2 | 4.96450 | 0.00713 |
| intent_fv_t1:fam | 741.3004 | 1 | 2.71762 | 0.09951 |
| frame:fam | 3251.1004 | 2 | 5.95930 | 0.00266 |
| intent_fv_t1:frame:fam | 3372.2251 | 2 | 6.18133 | 0.00213 |
| Residuals | 325147.9905 | 1192 | NA | NA |

| | eta.sq | eta.sq.part |
|---|---|---|
| intent_fv_t1 | 0.51192 | 0.59036 |
| frame | 0.00497 | 0.01379 |
| fam | 0.00027 | 0.00075 |
| intent_fv_t1:frame | 0.00296 | 0.00826 |
| intent_fv_t1:fam | 0.00081 | 0.00227 |
| frame:fam | 0.00355 | 0.00990 |
| intent_fv_t1:frame:fam | 0.00368 | 0.01026 |

#### Contrasts

| | Sum Sq | Df | F value | Pr(>F) |
|---|---|---|---|---|
| (Intercept) | 40226.2838 | 1 | 147.47048 | 0.00000 |
| intent_fv_t1 | 468588.8340 | 1 | 1717.85743 | 0.00000 |
| fam | 243.7821 | 1 | 0.89371 | 0.34467 |
| cont1 | 3154.3290 | 1 | 11.56384 | 0.00069 |
| cont2 | 1275.5733 | 1 | 4.67628 | 0.03078 |
| intent_fv_t1:fam | 741.3004 | 1 | 2.71762 | 0.09951 |
| intent_fv_t1:cont1 | 2460.5520 | 1 | 9.02044 | 0.00273 |
| fam:cont1 | 2429.1340 | 1 | 8.90526 | 0.00290 |
| intent_fv_t1:cont2 | 212.6870 | 1 | 0.77972 | 0.37741 |
| fam:cont2 | 902.9259 | 1 | 3.31015 | 0.06910 |
| intent_fv_t1:fam:cont1 | 2016.0481 | 1 | 7.39088 | 0.00665 |
| intent_fv_t1:fam:cont2 | 1433.4965 | 1 | 5.25523 | 0.02205 |
| Residuals | 325147.9905 | 1192 | NA | NA |

| | eta.sq | eta.sq.part |
|---|---|---|
| intent_fv_t1 | 0.51192 | 0.59036 |
| fam | 0.00027 | 0.00075 |
| cont1 | 0.00345 | 0.00961 |
| cont2 | 0.00139 | 0.00391 |
| intent_fv_t1:fam | 0.00081 | 0.00227 |
| intent_fv_t1:cont1 | 0.00269 | 0.00751 |
| fam:cont1 | 0.00265 | 0.00742 |
| intent_fv_t1:cont2 | 0.00023 | 0.00065 |
| fam:cont2 | 0.00099 | 0.00277 |
| intent_fv_t1:fam:cont1 | 0.00220 | 0.00616 |
| intent_fv_t1:fam:cont2 | 0.00157 | 0.00439 |

#### Group Means

| frame | emmean | SE | df | lower.CL | upper.CL |
|---|---|---|---|---|---|
| control | 71.16400 | 0.8733309 | 1192 | 69.45056 | 72.87743 |
| negative | 70.66860 | 0.9035879 | 1192 | 68.89580 | 72.44140 |
| positive | 75.66406 | 0.8805810 | 1192 | 73.93640 | 77.39172 |

| frame | fam | emmean | SE | df | lower.CL | upper.CL |
|---|---|---|---|---|---|---|
| control | unfamiliar | 69.34017 | 1.226393 | 1192 | 66.93404 | 71.74630 |
| negative | unfamiliar | 69.90138 | 1.246397 | 1192 | 67.45600 | 72.34675 |
| positive | unfamiliar | 75.84486 | 1.228014 | 1192 | 73.43555 | 78.25417 |
| control | familiar | 72.98782 | 1.243699 | 1192 | 70.54774 | 75.42790 |
| negative | familiar | 71.43581 | 1.308579 | 1192 | 68.86844 | 74.00319 |
| positive | familiar | 75.48326 | 1.262408 | 1192 | 73.00647 | 77.96005 |

### S1.7: Supplementary Model Removing Underestimators

More participants were found to underestimate side effects in the Unfamiliar group. The present analysis removes all those who underestimated side effects to determine whether their presence was responsible for driving differences between the Familiar and Unfamiliar vaccine types.

A similar pattern of results was observed to the primary model.

#### Primary Analysis: over-estimators only

#### Overall effects

| | Sum Sq | Df | F value | Pr(>F) |
|---|---|---|---|---|
| (Intercept) | 36182.6079 | 1 | 136.9021 | 0.0000 |
| intent_fv_t1 | 404453.3591 | 1 | 1530.3074 | 0.0000 |
| frame | 4023.0949 | 2 | 7.6110 | 0.0005 |
| fam | 124.1591 | 1 | 0.4698 | 0.4932 |
| intent_fv_t1:frame | 2337.6936 | 2 | 4.4225 | 0.0122 |
| intent_fv_t1:fam | 298.3073 | 1 | 1.1287 | 0.2883 |
| frame:fam | 3261.9273 | 2 | 6.1710 | 0.0022 |
| intent_fv_t1:frame:fam | 2531.4747 | 2 | 4.7891 | 0.0085 |
| Residuals | 266674.1528 | 1009 | NA | NA |

| | eta.sq | eta.sq.part |
|---|---|---|
| intent_fv_t1 | 0.5109 | 0.6026 |
| frame | 0.0051 | 0.0149 |
| fam | 0.0002 | 0.0005 |
| intent_fv_t1:frame | 0.0030 | 0.0087 |
| intent_fv_t1:fam | 0.0004 | 0.0011 |
| frame:fam | 0.0041 | 0.0121 |
| intent_fv_t1:frame:fam | 0.0032 | 0.0094 |

#### Contrasts

| | Sum Sq | Df | F value | Pr(>F) |
|---|---|---|---|---|
| (Intercept) | 36182.6079 | 1 | 136.9021 | 0.0000 |
| intent_fv_t1 | 404453.3591 | 1 | 1530.3074 | 0.0000 |
| fam | 124.1591 | 1 | 0.4698 | 0.4932 |
| cont1 | 2840.0663 | 1 | 10.7458 | 0.0011 |
| cont2 | 1135.9270 | 1 | 4.2979 | 0.0384 |
| intent_fv_t1:fam | 298.3073 | 1 | 1.1287 | 0.2883 |
| intent_fv_t1:cont1 | 1991.0113 | 1 | 7.5333 | 0.0062 |
| fam:cont1 | 2056.2021 | 1 | 7.7799 | 0.0054 |
| intent_fv_t1:cont2 | 319.0970 | 1 | 1.2073 | 0.2721 |
| fam:cont2 | 1246.5656 | 1 | 4.7166 | 0.0301 |
| intent_fv_t1:fam:cont1 | 1373.0765 | 1 | 5.1952 | 0.0229 |
| intent_fv_t1:fam:cont2 | 1201.0458 | 1 | 4.5443 | 0.0333 |

| | Sum Sq | Df | F value | Pr(>F) |
|---|---|---|---|---|
| Residuals | 266674.1528 | 1009 | NA | NA |

| | eta.sq | eta.sq.part |
|---|---|---|
| intent_fv_t1 | 0.5109 | 0.6026 |
| fam | 0.0002 | 0.0005 |
| cont1 | 0.0036 | 0.0105 |
| cont2 | 0.0014 | 0.0042 |
| intent_fv_t1:fam | 0.0004 | 0.0011 |
| intent_fv_t1:cont1 | 0.0025 | 0.0074 |
| fam:cont1 | 0.0026 | 0.0077 |
| intent_fv_t1:cont2 | 0.0004 | 0.0012 |
| fam:cont2 | 0.0016 | 0.0047 |
| intent_fv_t1:fam:cont1 | 0.0017 | 0.0051 |
| intent_fv_t1:fam:cont2 | 0.0015 | 0.0045 |

#### Group Means

| frame | fam | emmean | SE | df | lower.CL | upper.CL |
|---|---|---|---|---|---|---|
| control | unfamiliar | 68.09534 | 1.191541 | 1009 | 65.75716 | 70.43353 |
| negative | unfamiliar | 70.93460 | 1.576204 | 1009 | 67.84158 | 74.02761 |
| positive | unfamiliar | 74.25544 | 1.476670 | 1009 | 71.35774 | 77.15313 |
| control | familiar | 71.85229 | 1.247961 | 1009 | 69.40339 | 74.30119 |
| negative | familiar | 69.71158 | 1.377433 | 1009 | 67.00861 | 72.41454 |
| positive | familiar | 74.38450 | 1.356049 | 1009 | 71.72350 | 77.04550 |

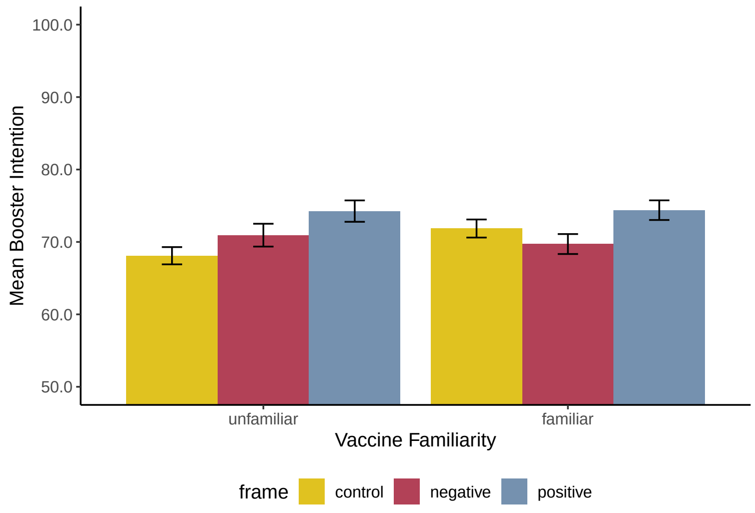


Figure 1.8.1: Graph depicting the Framing * Familiarity interaction among participants who did not underestimate side effects.

#### Means

#### Contrast Means

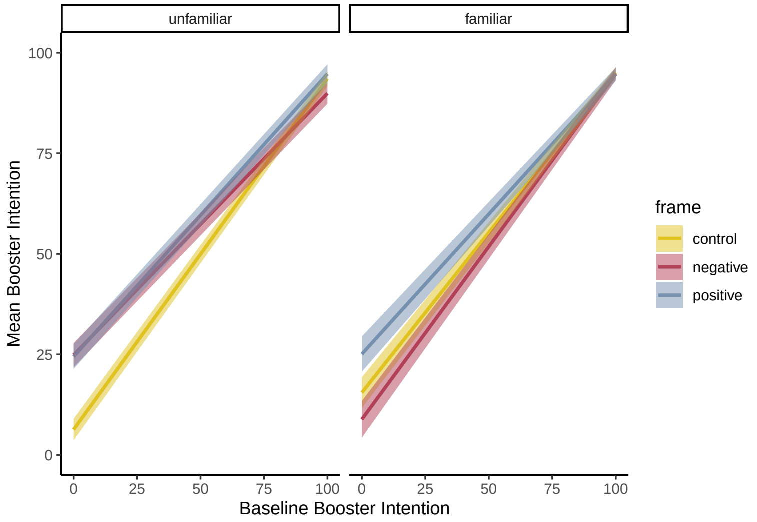


Figure 1.8.2: Graph depicting the Framing * Familiarity * Baseline Intention interaction among those participants who did not underestimate side effects.
